# Prevalence of chlamydia, gonorrhoea, and trichomoniasis among male and female general populations in sub-Saharan Africa from 2000-2024: A systematic review and meta-regression analysis

**DOI:** 10.1101/2024.12.16.24319070

**Authors:** Julia Michalow, Lauren Hall, Jane Rowley, Rebecca L. Anderson, Quinton Hayre, R. Matthew Chico, Olanrewaju Edun, Jesse Knight, Salome Kuchukhidze, Evidence Majaya, Domonique M. Reed, Oliver Stevens, Magdalene K Walters, Remco PH Peters, Anne Cori, Marie-Claude Boily, Jeffrey W. Imai-Eaton

## Abstract

**Background:** Sub-Saharan Africa (SSA) has the highest sexually transmitted infection (STI) prevalence globally, but information about trends and geographic variation is limited by sparse aetiologic studies, particularly among men. This systematic review assessed chlamydia, gonorrhoea, and trichomoniasis prevalence by sex, sub-region, and year, and estimated male-to-female prevalence ratios for SSA.

**Methods:** We searched Embase, MEDLINE, Global Health, PubMed, and African Index Medicus for studies measuring STI prevalence among general populations from January 1, 2000, to September 17, 2024. We adjusted observations for diagnostic test performance and used log-binomial mixed-effects meta-regressions to estimate prevalence trends and sex-prevalence ratios.

**Findings:** Of 5202 records identified, we included 211 studies from 28 countries. In 2020, estimated prevalence among 15-49-year-olds in SSA for chlamydia was 6.6% (95%CI: 5.0-8.2%, n=169 observations) among females and 4.7% (3.4-6.1%, n=33) among males, gonorrhoea was 2.4% (1.4-3.3%, n=171) and 1.7% (0.7-2.6%, n=31), and trichomoniasis was 6.8% (3.6-9.9%, n=188) and 1.7% (0.7-2.7%, n=19). Male-to-female ratio estimates were 0.61 (0.53-0.71) for chlamydia, 0.81 (0.61-1.09) for gonorrhoea, and 0.23 (0.18-0.28) for trichomoniasis. From 2010-2020, chlamydia prevalence increased by 34.5% (11.1-62.9%) in SSA, while gonorrhoea and trichomoniasis trends were not statistically significant. Chlamydia and gonorrhoea prevalence were highest in Southern and Eastern Africa, whereas trichomoniasis was similar across sub-regions.

**Interpretation:** SSA has a high, geographically varied STI burden, with increasing prevalence of chlamydia. Region-specific sex-prevalence ratios differed from existing global ratios and should be considered in future burden estimates. Enhanced sex-stratified surveillance is crucial to guide national programmes and reduce STI prevalence in SSA.

**Funding:** Gates Foundation, Imperial College London, NIH, UKRI

## Background

The World Health Organization (WHO) has established targets to reduce the incidence of sexually transmitted infections (STIs) by 2030.^1^ Achieving these targets requires ambitious scale-up of national STI prevention and control programmes, particularly in sub-Saharan Africa (SSA) where STI prevalence is highest globally.^1^ However, monitoring and optimising these programmes is challenging due to high rates of asymptomatic infection, variable health-seeking behaviour, limited diagnostic capacity necessitating syndromic management, and weak surveillance systems.^2^ STI prevalence estimates are crucial for evaluating progress toward WHO targets but are hindered by infrequent population-representative studies, especially among men.^3^

Estimates of curable STI prevalence in SSA are limited. WHO publishes global and regional prevalence and incidence estimates for curable STIs, including chlamydia (*Chlamydia trachomatis*, CT), gonorrhoea (*Neisseria gonorrhoeae*, NG), and trichomoniasis (*Trichomonas vaginalis*, TV), approximately every four years.^4–9^ These focus on global geographic regions and do not report sub-regional variation or temporal trends. The Global Burden of Disease (GBD) study generates global, regional, and national prevalence and incidence trends for several STIs.^10^ Both estimation approaches depend heavily on available data and underlying model assumptions. Due to limited data among men, WHO and GBD have derived male-to-female prevalence ratios from global data and applied these in most regions to support male prevalence estimates.^5,8,10^ Although WHO reviews its global ratios prior to each estimation round, values have not been updated since 1999 for trichomoniasis and 2005 for chlamydia and gonorrhoea. ^5,8^ Additionally, several systematic reviews have assessed the prevalence of curable STIs in SSA sub-regions among pregnant women,^11,12^ women participating in HIV prevention trials,^13^ or women living with HIV,^14^ but their generalisability beyond the specified populations is limited and these reviews have not examined changes in STI prevalence over time.

We therefore conducted a systematic review of studies on chlamydia, gonorrhoea, and trichomoniasis prevalence among adults considered representative of the general population (i.e. not conducted among groups identified as key populations, as defined in WHO guidelines on HIV, STIs, and viral hepatitis^1^) in SSA from 2000 to 2024. We performed meta-analyses to assess variations in STI prevalence by sex, sub-region, and over time. To inform future regional and global STI burden estimation efforts, we derived updated male-to-female prevalence ratios specific to SSA.

## Methods

### Search strategy

We systematically searched Embase (Ovid), MEDLINE (Ovid), Global Health (Ovid), PubMed, and African Index Medicus from 1 January 2000 to 17 September 2024 for studies assessing the prevalence of chlamydia, gonorrhoea, and trichomoniasis in SSA. Search term domains included relevant terms and synonyms for “sexually transmitted infections” and “sub-Saharan Africa” (Table S1). We also searched citations from relevant published systematic reviews.^7–9,11–20^

### Eligibility criteria and study selection

We included studies reporting sex-stratified empirical data on the prevalence of chlamydia, gonorrhoea, or trichomoniasis among adults from the general population in SSA between 2000 and 2024. We considered studies to represent the general population if they enrolled participants from antenatal care (ANC), family planning, or gynaecology clinics; primary healthcare or other outpatient services; educational institutions; or community venues (e.g., social spaces or places of worship); or recruited participants for HIV or STI prevention trials (baseline data only) or population-representative surveys. We excluded studies if participants sought treatment for STI symptoms or infertility or were enrolled exclusively from populations living with HIV or key populations.

We included studies with participants aged 12 years or older who were tested using internationally recognised aetiologic diagnostic tests with urogenital specimens (urine, urethral, or cervicovaginal samples), and excluded studies that only collected pharyngeal or rectal samples. We also excluded case reports, commentaries, longitudinal and randomised controlled studies reporting only post-baseline outcomes, and studies with fewer than 15 participants. We used the UN M49 standard to define SSA and its sub-regions (Table S2).^21^

Search results were managed and de-duplicated using Covidence systematic review software (Veritas Health Innovation, Melbourne, Australia).^22^ Researchers (DMR, EM, JM, JWI-E, JK, LH, MKW, OE, OS, QH, RLA, SK) independently double-screened title and abstract records for eligibility and assessed full text articles for inclusion. A third reviewer resolved discrepancies.

### Data extraction

Researchers (JM, LH, RLA, QH) independently double-extracted prevalence observations (number of individuals with a positive result of the total number tested) for each study with stratification by country, sex, and population group, as available. We also recorded information on study characteristics, participant characteristics, and diagnostic methodology (Table S3). Discrepancies were resolved through consensus.

If outcomes from the same study were reported in multiple articles, we preferentially extracted observations from the largest sample or, if samples were the same size, the first published article. For studies collecting samples from multiple anatomical sites, we extracted only urogenital results. If multiple diagnostic tests were conducted, we extracted outcomes from the test and sample type with highest sensitivity and specificity for pathogen identification (Table S4). We preferentially extracted non-stratified rather than HIV-stratified outcomes, if both were reported.

### Data analysis

We adjusted study prevalence estimates to account for imperfect diagnostic test sensitivity and specificity, using a previously described Bayesian approach.^23^ Sensitivity and specificity values for each test category were collated from published literature, with preference for characteristics published by the WHO (Table S4).^5,23,24^

For studies that did not report data collection dates (n=26), we imputed the midpoint year of data collection by subtracting the median publication lag from the publication year (three years, based on the difference between the data collection midpoint year and year of publication among studies with dates reported). For one study spanning multiple sub-regions without country- or region-specific data reported,^25^ we classified the study sub-region according to where most participants had been recruited.

To estimate sub-regional trends in sex-specific STI prevalence in SSA, we fit log-binomial mixed-effects meta-regressions for each pathogen. Fixed effects included year (midpoint of data collection), study population (ANC attendees, family planning clinic attendees, gynaecology clinic attendees, primary healthcare or other outpatient facility attendees, students, community members recruited from non-clinical venues, HIV/STI prevention trial participants, and population-representative survey participants), sex (female, male), age group (younger adults 12– 25 years, all adults ≥ 12 years), HIV status (HIV negative, non-stratified HIV status), diagnostic test (nucleic acid amplification test [NAAT], culture, direct fluorescent antibody, enzyme immunosorbent assay, rapid antigen test, wet mount), sub-region (Western and Central Africa [WCA], Eastern Africa [EA], or Southern Africa [SA]), year and sub-region interaction term, and sex and sub-region interaction term. A year and sex interaction term was not included due to limited observations of male prevalence. We combined Western and Central Africa due to limited prevalence observations in Central Africa. Study-level random intercepts allowed for between-study heterogeneity. Fitted models were used to predict the prevalence of each infection among male and female adults by sub-region between 2000 and 2024, using reference categories for covariates in the model (study population: ANC attendees, HIV status: non-stratified, diagnostic test: NAAT). We report estimates for sub-regions and all of SSA for 2020. SSA estimates were obtained by weighting sub-regional predictions with sex-matched population size for adults aged 15-49 years in 2020, according to UN World Population Prospects 2024.^26^ Standard errors were determined using the delta method.^27^

To assess sex differences in prevalence, we estimated male-to-female prevalence ratios using observations from all studies (between-study ratios) and for the subset of studies providing estimates for both sexes (within-study ratios). We estimated between-study ratios using previously described meta-regression models. We determined within-study ratios using the same meta-regression approach, but without fixed effects for population group and an interaction term for sex and sub-region, due to limited observations. We also derived pooled within-study ratios without adjusting for any covariates.

We assessed sensitivity of prevalence estimates, trends, and male-to-female prevalence-ratios to adjustments for diagnostic performance by comparing results from models using all available observations versus only NAAT-diagnosed observations, with and without accounting for test sensitivity and specificity. Unless indicated otherwise, all main results reflect observations adjusted for diagnostic test performance. Results are presented as means with 95% confidence intervals (95% CIs). Model coefficients are presented as adjusted prevalence ratios (aPRs), with confidence intervals calculated on the log scale prior to exponentiation. Analyses were conducted in R version 4.2.3; Bayesian diagnostic test performance adjustments used rstan version 2.26.21^28^ and meta-regression modelling used glmmTMB version 1.1.8.^29^

We pre-registered our study with PROSPERO (CRD: 42023420384)^30^ and reported according to the Preferred Reporting Items for Systematic Reviews and Meta-Analyses (PRISMA) guideline.^31^ The Imperial College Research Ethics Committee approved analysis of secondary data for this study (ICREC #6365329).

## Results

### Search results and scope

We identified 11,136 records through the database search, of which 5,934 were duplicates and 5,202 were screened (Figure 1). Of these, 783 full-text articles were assessed and 301 met eligibility criteria. After adding 11 articles identified through citation searching and excluding 98 articles with duplicate study observations, we included 214 articles from 211 unique studies, from which we extracted 614 prevalence observations (NG=203, CT=203, TV=208; Table S5). Most studies were in Eastern Africa (N=90/211, 42.7%), followed by Western and Central (N=71/211, 33.6%), and Southern (N=59/211, 28.0%) Africa (Table 1). Studies were conducted in 28 different sub-Saharan African countries, with most in South Africa (N=53/211, 25.1%), Nigeria (N=38/211, 18.0%), Kenya (N=26/211, 12.3%), Tanzania (N=20/211, 9.5%), and Uganda (N=19/211, 9.0%) (Figure S1). Study populations were predominantly ANC attendees (N=85/211, 40.3%), primary healthcare or outpatient department attendees (N=30/211, 14.2%), and HIV/STI prevention trial participants (N=24/211, 11.4%). Few studies were among population-representative survey samples (N=11/211, 5.2%). Most studies were solely among females (N=174/211, 82.5%). A limited number included both sexes (N=29/211, 13.7%) or males only (N=8/211, 3.8%). Studies were predominantly among all adults (N=180/211, 85.3%), rather than restricted to younger adults (N=31/211, 14.6%). Across all studies, most enrolled within the 15-49-year age range (N=109/211, 51.7%), while others included younger and/or older participants (N=45/211, 21.3%), reported only a minimum adult age (N=37/211, 17.5%), or did not report age ranges (N=20/211, 9.5%).

**Figure 1:**
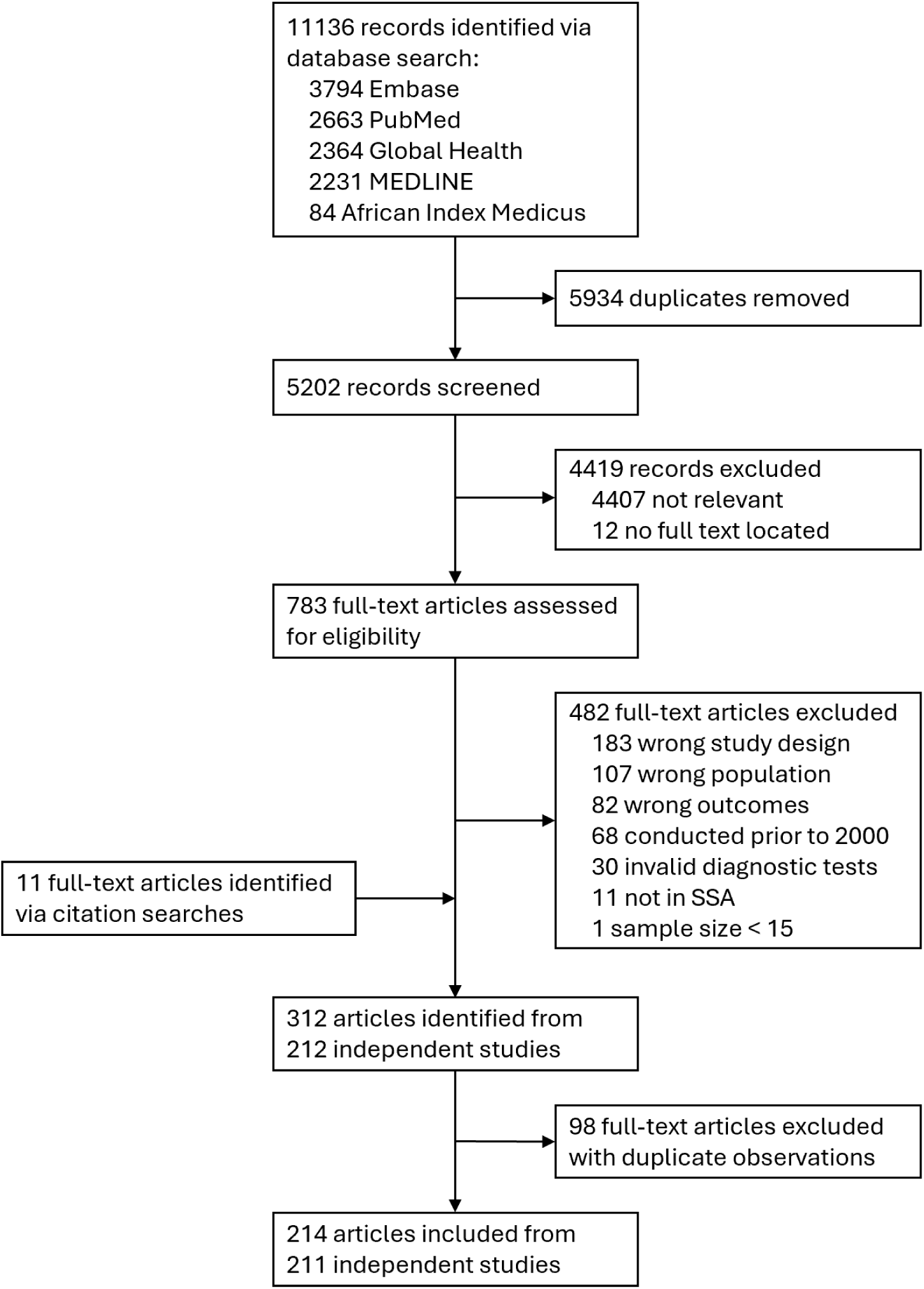
Study selection flowchart. Flowchart of study selection for systematic review.

**Table 1:** Summary of characteristics for included studies.

| Variable | Group | Chlamydia<br>(N=139) | Gonorrhoea<br>(N=140) | Trichomoniasis<br>(N=162) | Total<br>(N=211) |
| --- | --- | --- | --- | --- | --- |
| **Sub-region<sup>1</sup>** | Western Africa | 20 (14.4%) | 20 (14.3%) | 51 (31.5%) | 59 (28.0%) |
|  | Central Africa<sup>2</sup> | 10 (7.2%) | 8 (5.7%) | 8 (4.9%) | 12 (5.7%) |
|  | Eastern Africa | 69 (49.6%) | 70 (50.0%) | 63 (38.9%) | 90 (42.7%) |
|  | Southern Africa | 49 (35.3%) | 51 (36.4%) | 46 (28.4%) | 59 (28.0%) |
|  | Multiple sub-regions<sup>3</sup> | 1 (0.7%) | 1 (0.7%) | 1 (0.6%) | 1 (0.5%) |
| **Study**<br>**midpoint**<br>**year<sup>1,4</sup>** | 2000-2004 | 27 (19.4%) | 32 (22.9%) | 34 (21.0%) | 45 (21.3%) |
|  | 2005-2009 | 18 (12.9%) | 19 (13.6%) | 21 (13.0%) | 27 (12.8%) |
|  | 2010-2014 | 35 (25.2%) | 31 (22.1%) | 38 (23.5%) | 51 (24.2%) |
|  | 2015-2019 | 42 (30.2%) | 41 (29.3%) | 54 (33.3%) | 64 (30.3%) |
|  | 2020-2024 | 18 (12.9%) | 17 (12.1%) | 15 (9.3%) | 25 (11.8%) |
| **Population**<br>**group<sup>1</sup>** | ANC attendees | 39 (28.1%) | 42 (30.0%) | 75 (46.3%) | 85 (40.3%) |
|  | FP attendees | 12 (8.6%) | 12 (8.6%) | 11 (6.8%) | 17 (8.1%) |
|  | GYN attendees | 6 (4.3%) | 8 (5.7%) | 11 (6.8%) | 14 (6.6%) |
|  | PHC/OPD attendees | 22 (15.8%) | 22 (15.7%) | 20 (12.3%) | 30 (14.2%) |
|  | Students | 15 (10.8%) | 11 (7.9%) | 15 (9.3%) | 19 (9.0%) |
|  | Community members<sup>5</sup> | 15 (10.8%) | 16 (11.4%) | 6 (3.7%) | 17 (8.1%) |
|  | HIV/STI prevention trial<br>participants<sup>6</sup> | 22 (15.8%) | 22 (15.7%) | 22 (13.6%) | 24 (11.4%) |
|  | Population-representative<br>survey participants | 10 (7.2%) | 10 (7.1%) | 8 (4.9%) | 11 (5.2%) |
| **Sex** | Female only | 106 (76.3%) | 109 (77.9%) | 143 (88.3%) | 174 (82.5%) |
|  | Male only | 7 (5.0%) | 5 (3.6%) | 7 (4.3%) | 8 (3.8%) |
|  | Both sexes (stratified) | 26 (18.7%) | 26 (18.6%) | 12 (7.4%) | 29 (13.7%) |
| **Age group** | Adult (12+ years) | 112 (80.6%) | 116 (82.9%) | 140 (86.4%) | 180 (85.3%) |
|  | Youth (12–25 years) | 27 (19.4%) | 25 (17.9%) | 22 (13.6%) | 31 (14.7%) |
| **HIV status<sup>1,7</sup>** | HIV-negative | 37 (26.6%) | 38 (27.1%) | 38 (23.5%) | 46 (21.8%) |
|  | Non-stratified | 103 (74.1%) | 103 (73.6%) | 125 (77.2%) | 166 (78.7%) |

N: number of studies, ANC: antenatal care, FP: family planning, GYN: gynaecology clinic, PHC/OPD: Primary healthcare or outpatient department.
<sup>1</sup>The same study is included in more than one subcategory if it reports across different variable levels or when multiple levels are relevant. <sup>2</sup> Western and Central Africa combined for analysis. <sup>3</sup>Study without country- or region-specific reporting was allocated to Eastern Africa for analysis, according to where most participants were recruited. <sup>4</sup>Midpoint year between start and end of data collection period. <sup>5</sup>Community members recruited through non-clinical venues, such as social spaces or places of worship. <sup>6</sup>Studies reporting baseline data among trial participants. <sup>7</sup>Studies exclusively among populations living with HIV were excluded. Data were preferentially extracted and analysed without stratifying by HIV status if both were reported.

### Sexually transmitted infection prevalence

In 2020, estimated prevalence of chlamydia in SSA was 6.6% (95%CI: 5.0-8.2%, number of observations (n)=169) among females and 4.7% (3.4-6.1%, n=33) among males, gonorrhoea was 2.4% (1.4-3.3%, n=171) and 1.7% (0.7-2.6%, n=31), and trichomoniasis was 6.8% (3.6-9.9%, n=188) and 1.7% (0.7-2.7%, n=19), respectively (Figure 2A). From 2010 to 2020, chlamydia prevalence increased by 34.5% (11.1-62.9%) in SSA, while trends for gonorrhoea (3.3% increase [-20.2-33.9%]) and trichomoniasis (17.8% increase [-3.8-34.9%]) were not statistically significant (Table 2, Figure 2B).

**Figure 2:**
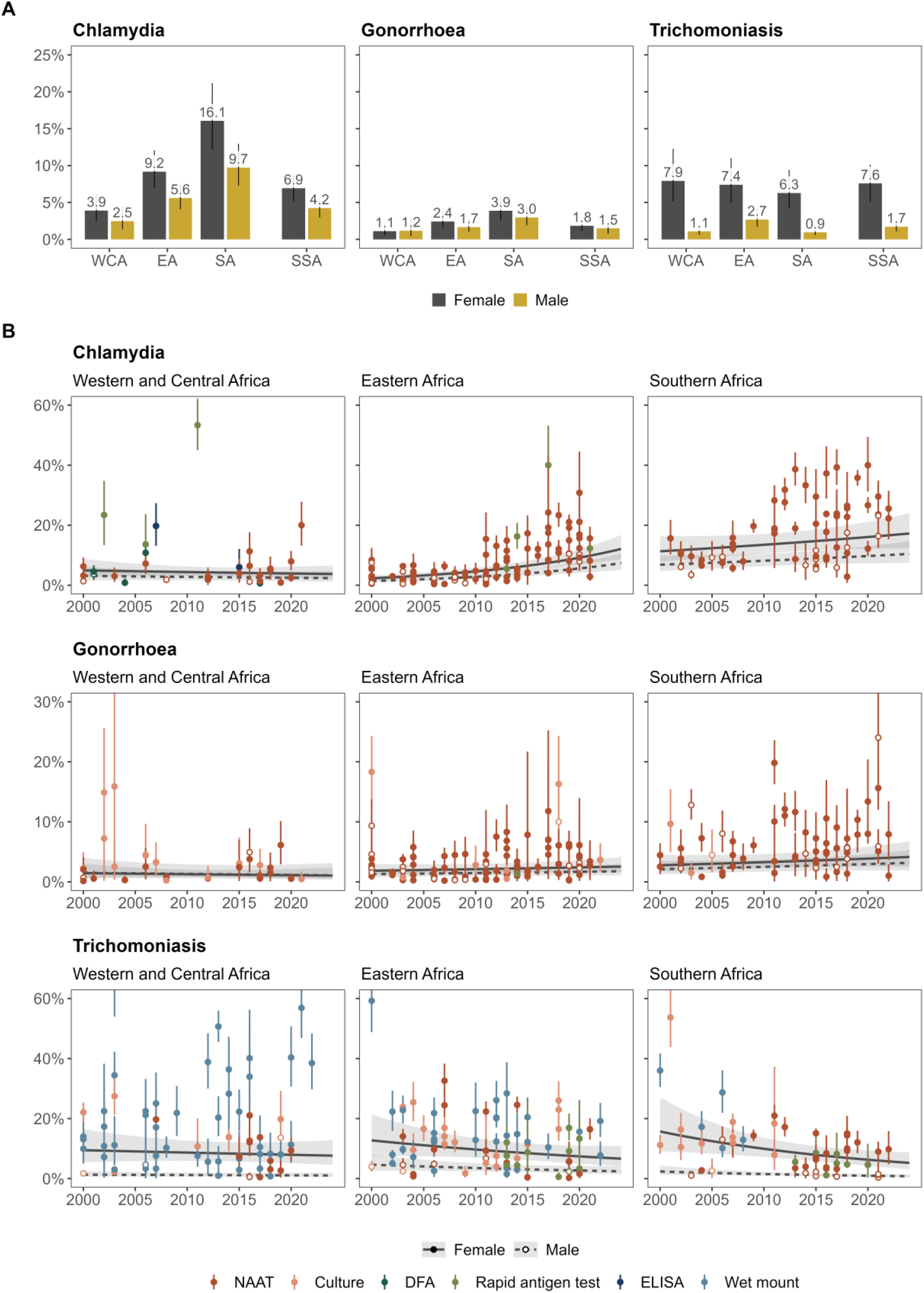
Sexually transmitted infection prevalence per sub-region in sub-Saharan Africa. Estimates of chlamydia, gonorrhoea, and trichomoniasis prevalence by sex and sub-region. Sub-regional estimates generated using log-binomial generalised linear mixed-effects models for each infection. Sub-Saharan African estimates represent sex-matched population-weighted means. <u>Panel A</u>: Prevalence estimates in 2020. Bars and error lines depict mean prevalence estimates with 95% confidence intervals. <u>Panel B</u>: Prevalence estimates between 2000 and 2024. Lines and shaded areas depict mean prevalence estimates (solid line: females, dashed line: males) with 95% confidence intervals. Points and error lines represent study prevalence observations (solid point: female, open point: male) with 95% confidence intervals, colour coded according to diagnostic test category. Y-axes truncated. DFA: Direct fluorescent antibody, EA: Eastern Africa, NAAT: nucleic acid amplification test, SA: Southern Africa, SSA: Sub-Saharan Africa, WCA: Western and Central Africa.

**Table 2:** Adjusted prevalence ratios for chlamydia, gonorrhoea, and trichomoniasis in sub-Saharan Africa, estimated via log-binomial generalised linear mixed-effects models.

| Variable | Chlamydia<br>aPR (95% CI) | Gonorrhoea<br>aPR (95% CI) | Trichomoniasis<br>aPR (95% CI) |
| --- | --- | --- | --- |
| **Intercept** | 0.14 (0.11-0.18) | 0.03 (0.02-0.05) | 0.09 (0.07-0.12) |
| **Sub-region** |  |  |  |
| Western and Central | 0.31 (0.22-0.43) | 0.36 (0.23-0.57) | 0.94 (0.68-1.31) |
| Eastern | 0.38 (0.35-0.41) | 0.64 (0.56-0.74) | 1.01 (0.85-1.21) |
| Southern | Ref | Ref | Ref |
| **Sub-region:Sex<sup>1</sup>** |  |  |  |
| Western and Central:Male | 0.63 (0.43-0.93) | 1.05 (0.54-2.04) | 0.14 (0.09-0.20) |
| Eastern:Male | 0.61 (0.53-0.70) | 0.69 (0.56-0.85) | 0.36 (0.30-0.44) |
| Southern:Male | 0.61 (0.55-0.66) | 0.76 (0.65-0.90) | 0.15 (0.11-0.20) |
| **Sub-region:Year<sup>2</sup>** |  |  |  |
| Western and Central:Year | 0.99 (0.95-1.03) | 0.99 (0.94-1.04) | 0.99 (0.96-1.02) |
| Eastern:Year | 1.07 (1.05-1.09) | 1.01 (0.99-1.04) | 0.97 (0.94-1.01) |
| Southern:Year | 1.02 (1.00-1.04) | 1.02 (0.99-1.05) | 0.96 (0.92-0.99) |
| **Population** |  |  |  |
| ANC attendees | Ref | Ref | Ref |
| FP attendees | 0.95 (0.63-1.45) | 0.90 (0.50-1.64) | 0.54 (0.32-0.90) |
| GYN attendees | 1.03 (0.57-1.88) | 0.87 (0.42-1.82) | 1.34 (0.80-2.25) |
| PHC/OPD attendees | 0.81 (0.57-1.14) | 1.49 (0.92-2.39) | 0.94 (0.61-1.43) |
| Students | 1.06 (0.69-1.63) | 1.14 (0.56-2.31) | 1.23 (0.76-1.99) |
| Community members | 0.97 (0.66-1.44) | 1.10 (0.65-1.87) | 0.96 (0.50-1.86) |
| HIV/STI prevention trial participants | 0.89 (0.61-1.28) | 1.20 (0.72-1.99) | 1.00 (0.62-1.63) |
| Population-representative survey participants | 0.90 (0.58-1.42) | 1.19 (0.64-2.20) | 1.32 (0.71-2.44) |
| **Age group** |  |  |  |
| All adults (12+ years) | Ref | Ref | Ref |
| Younger adults (12 - 25 years) | 1.14 (0.85-1.52) | 1.12 (0.75-1.68) | 0.55 (0.35-0.88) |
| **HIV status** |  |  |  |
| Non-stratified | Ref | Ref | Ref |
| HIV negative | 1.42 (1.06-1.90) | 1.06 (0.71-1.59) | 0.79 (0.56-1.14) |
| **Diagnostic test** |  |  |  |
| NAAT | Ref | Ref | Ref |
| Culture | - | 1.60 (1.00-2.54) | 1.36 (0.89-2.09) |
| DFA | 0.70 (0.32-1.51) | - | - |
| ELISA | 2.26 (0.86-5.98) | - | - |
| Rapid antigen test | 2.94 (1.76-4.92) | 0.52 (0.06-4.61) | 1.03 (0.56-1.90) |
| Wet mount | - | - | 1.61 (1.14-2.29) |
| **Model variance** |  |  |  |
| $\tau^2$ fixed | 0.51 | 0.16 | 0.45 |
| $\tau^2$ random | 0.34 | 0.60 | 0.60 |
| **Number groups** |  |  |  |
| Number studies | 139 | 140 | 162 |
| Number observations | 202 | 202 | 207 |

Generalised linear mixed-effects models on the risk of chlamydia, gonorrhoea, and trichomoniasis in sub-Saharan Africa, using all study observations adjusted for diagnostic test performance.
<sup>1</sup>Reference sex is female. <sup>2</sup>Midpoint year between start and end of data collection period; centred at 2012.
aPR: adjusted prevalence ratio, 95% CI: 95% confidence interval, ANC: antenatal care, DFA: direct fluorescent antibody, FP: family planning clinic, GYN: gynaecology clinic, NAAT: nucleic acid amplification test, PHC/OPD: primary health care or outpatient department, Ref: Reference group, $\tau^2$ : variance.

Sub-regional variations in prevalence were similar for chlamydia and gonorrhoea (Table 2, Figure 2A). Among females, relative to Southern Africa, the prevalence of chlamydia and gonorrhoea were lowest in Western and Central Africa (aPR_CT_: 0.31 [0.22-0.43], aPR_NG_: 0.36 [0.23-0.57]), followed by Eastern Africa (aPR_CT_: 0.38 [0.35-0.41], aPR_NG_: 0.64 [0.56-0.74]). Similarly for males, prevalence was lower in Western and Central (aPR_CT_: 0.38 [0.32-0.46], aPR_NG_: 0.58 [0.44-0.78]) and Eastern Africa (aPR_CT_: 0.32 [0.19-0.53], aPR_NG_: 0.50 [0.23-1.10]) compared to Southern Africa. For trichomoniasis, female prevalence was similar across sub-regions (WCA aPR_TV_: 0.94 [0.68-1.31], EA aPR_TV_: 1.01 [0.85-1.21]), while male prevalence was higher in Eastern (aPR_TV_: 2.45 [1.66-3.61]) than Southern Africa.

Time trends for each infection varied across sub-regions. Chlamydia prevalence increased in Eastern (aPR_CT_ per year: 1.07 [1.05-1.09]) and Southern Africa (aPR_CT_: 1.02 [1.00-1.04]), while trichomoniasis prevalence decreased in Southern Africa (aPR_TV_: 0.96 [0.92-0.99]) only. Gonorrhoea prevalence was stable across all three sub-regions (Table 2, Figure 2B).

Across study populations, there was no evidence that the prevalence of any infection differed compared to ANC attendees, except for lower trichomoniasis prevalence among family planning clinic attendees (aPR: 0.54 [0.32-0.90]). Studies diagnosing chlamydia with a rapid antigen test (aPR_CT_: 2.94 [1.76-4.92]), gonorrhoea with culture (aPR_NG_: 1.60 [1.00-2.54]), and trichomoniasis with wet mount (aPR_TV_: 1.61 [1.14-2.29]) had higher prevalence than those that used NAAT, after accounting for diagnostic test performance (Table 2).

### Male-to-female prevalence ratio estimates

In SSA, male-to-female prevalence ratio estimates for each infection were similar when estimated using adjusted between-study ratios (all available data), adjusted within-study ratios (paired data), or unadjusted pooled within-study ratios (Figure 3, Table 2, Table S6). Male-to-female prevalence ratio estimates (adjusted between-study) for SSA in 2020 were 0.61 (0.53-0.71) for chlamydia, 0.81 (0.61-1.09) for gonorrhoea, and 0.23 (0.18-0.28) for trichomoniasis (Figure 3, Table 2). Between-study prevalence ratios were consistent across sub-regions for chlamydia (WCA:0.63 [0.43-0.93], EA:0.61 [0.53-0.70], SA:0.61 [0.55-0.66]), but varied for gonorrhoea (WCA:1.05 [0.54-2.04], EA:0.69 [0.56-0.85], SA:0.76 [0.65-0.90]) and trichomoniasis (WCA:0.14 [0.09-0.20], EA:0.36 [0.30-0.44], SA:0.15 [0.11-0.20]; Table 2).

**Figure 3:**
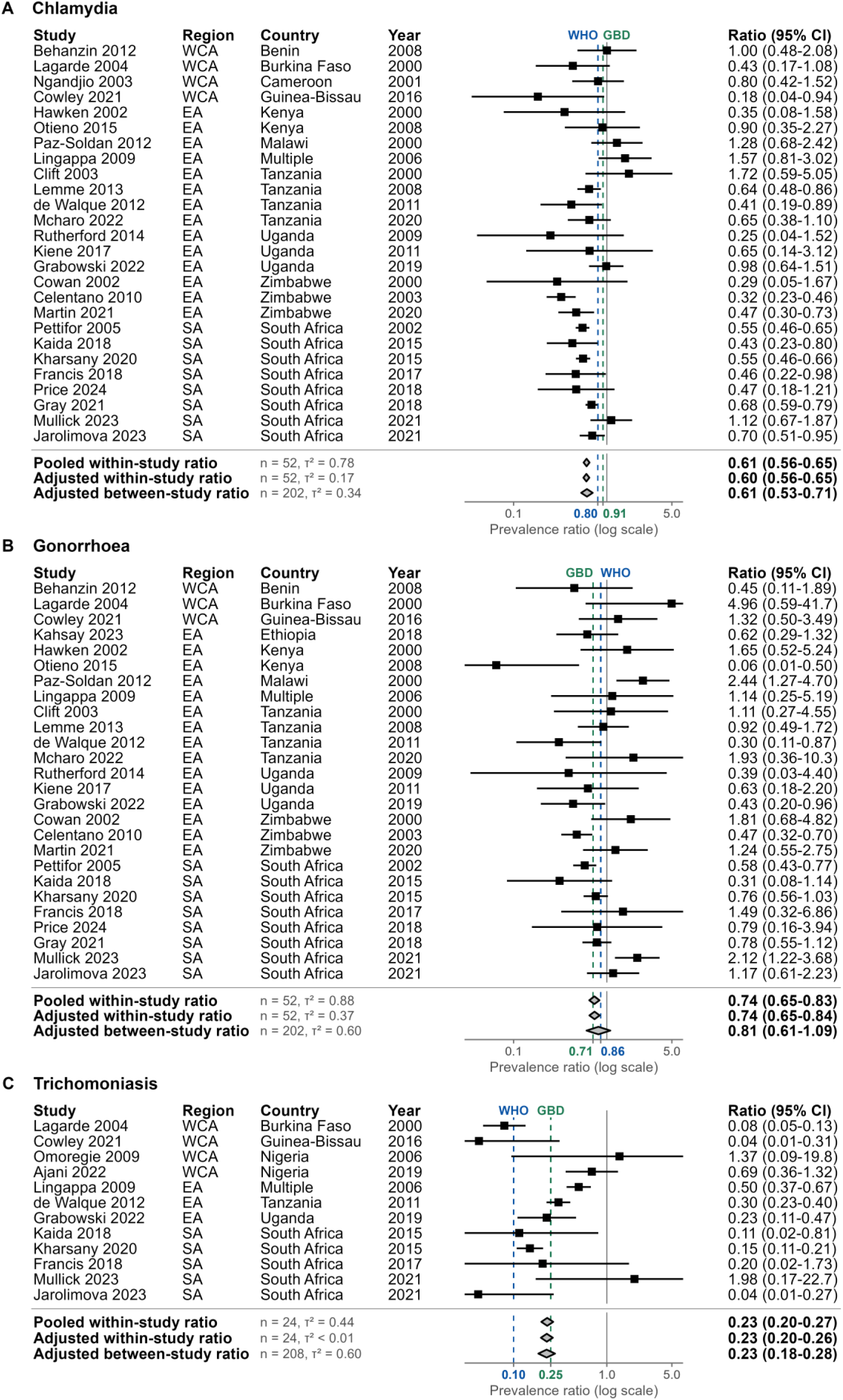
Sexually transmitted infection male-to-female prevalence ratio estimates in sub-Saharan Africa. Study observations and aggregate estimates of male-to-female prevalence ratios for (A) chlamydia, (B) gonorrhoea and (C) trichomoniasis. Ratios estimated using log-binomial generalised linear mixed-effects models per infection. Models for within-study ratios used study observations among both sexes and between-study ratios used all study observations. All models included sex as a fixed effect and study-level random intercepts. Pooled ratio models did not adjust for additional covariates. Adjusted ratio models included fixed effects for region, year, age-group, HIV-status, diagnostic test, and population group (between-study only). Points and error lines depict ratios and 95% confidence intervals for studies with observations among both sexes. Diamonds depict population-weighted mean ratios and 95% confidence intervals for sub-Saharan Africa, with adjusted ratios reported for 2020. Blue dotted lines and x-axis labels represent WHO global male-to-female prevalence ratios.^5,8^ Green dotted lines and x-axis labels represent the Global Burden of Disease study male-to-female prevalence ratios.^10^ X-axes are truncated. EA: Eastern Africa, SA: Southern Africa, WCA: Western and Central Africa, n: number of observations, τ^2^: variance for study random intercept.

### Sensitivity analyses of diagnostic test performance

Prevalence estimates in 2020 were consistent for all three infections when derived using all available or NAAT-only observations, with or without accounting for diagnostic test performance (Table 2, Table S7-S9, Figure S2). Temporal trends for chlamydia and gonorrhoea also remained consistent across sub-regions. However, for trichomoniasis, all-observation models showed a prevalence decline in Southern Africa (aPR: 0.96 [0.92-0.99] with and without test performance), while NAAT-only models did not (with test performance: 0.96 [0.90-1.02]; without: 0.96 [0.92-1.01]; Table 2, Table S7-S9, Figure S3). Between-study and within-study male-to-female prevalence ratios were consistent for all three infections, whether derived using all available or NAAT-only observations. However, accounting for test performance led to higher ratios for gonorrhoea and lower ratios for chlamydia and trichomoniasis (Table 2, Table S7-S9, Figure S4).

## Discussion

Our analysis estimated the prevalence of chlamydia, gonorrhoea and trichomoniasis among general populations in SSA between 2000 and 2024. We estimated a higher burden of curable STIs among females (6.9% chlamydia, 1.8% gonorrhoea, and 7.6% trichomoniasis in 2020) than males (4.2%, 1.5%, and 1.7% respectively), which exceeded global prevalence estimates (3.2%, 0.7%, and 2.7% for both sexes in 2020, respectively).^9^ Geographic variation was substantial; prevalence of chlamydia and gonorrhoea was highest in Southern Africa, followed by Eastern Africa and then Western and Central Africa. Trichomoniasis prevalence was similar across sub-regions for females but was highest in Eastern Africa for males. The prevalence of chlamydia increased considerably in Eastern and Southern Africa over the study period, while gonorrhoea was stable, and trichomoniasis decreased in Southern Africa.

Our central estimates of STI prevalence in 2020 differed from those reported by WHO for the African region and GBD for SSA.^9,10^ For females, our estimates were higher for chlamydia (6.9% vs. WHO:5.5%, GBD:4.5%), similar for gonorrhoea (1.8% vs. WHO:1.6%, GBD:1.7%), and lower for trichomoniasis (7.6% vs. WHO:12.0%, GBD:12.6%). For males, our estimates were higher for chlamydia (4.2% vs. WHO:4.0%, GBD:2.5%), gonorrhoea (1.5% vs. WHO:1.2%, GBD:1.2%), and trichomoniasis (1.7% vs. WHO:1.3%, GBD:1.5%).^9,10^ These differences reflect our broader study inclusion criteria, incorporation of more recent studies, and analysis of temporal trends to reflect sub-regional changes in prevalence. Compared to existing global male-to-female prevalence ratios, our SSA ratio estimates were lower for chlamydia (WHO: 0.80 and GBD: 0.91), and intermediate for gonorrhoea (WHO: 0.86 and GBD: 0.71) and trichomoniasis (WHO: 0.10 and GBD: 0.25; Figure 3).^5,8,10^ Although our analysis indicated sub-regional ratio variation, data limitations precluded more detailed exploration. Given these differences, future estimation exercises should prioritise using region-specific ratios to capture local epidemiologic differences, although sparse male prevalence data can hinder ratio estimation.

The spatial distribution of chlamydia and gonorrhoea prevalence mirror that of HIV prevalence in SSA, characterised by highest burden in Southern Africa, followed by Eastern Africa, and lowest in Western and Central Africa.^32^ This is likely due to shared behavioural risk factors and biological synergies that facilitate co-infection.^33^ Despite evidence suggesting that trichomoniasis increases HIV transmission and acquisition risk,^34^ prevalence variations were not observed across sub-regions. This could reflect limitations in study availability and diagnostics, missing covariates, or incomplete adjustment for covariates such as age.^35^ Greater integration of STI and HIV services remains important for their effective mitigation in high-burden areas.^1^

The increasing prevalence of chlamydia in Eastern and Southern Africa and decreasing prevalence of trichomoniasis in Southern Africa have not been documented previously. Previous STI burden assessments reported stable prevalence over time for chlamydia and gonorrhoea in Kenya and South Africa^36,37^ or did not estimate time trends for curable STIs in SSA.^8,11–14^ For chlamydia, increases in prevalence are particularly concerning given the severe, long-term health consequences of untreated infection, including chronic pelvic pain, infertility, ectopic pregnancy, and other adverse pregnancy outcomes, along with an increased risk of HIV transmission.^1^ For trichomoniasis, declines in prevalence warrant cautious interpretation. Our trend estimates were sensitive to the diagnostic tests included, due to more widespread use of NAAT rather than wet mount microscopy in recent studies.

Our analysis was primarily limited by sparse and heterogenous data. Our systematic search identified only 211 studies in 28 of 48 SSA countries. There were particularly few studies in Central Africa (12 studies in 6 countries) and among men (37 studies in 15 countries). We were therefore unable to generate estimates specific to Central Africa or to determine whether temporal prevalence trends differed by sex. Our estimates for Southern Africa relied predominantly on studies conducted in South Africa (51 of 59 studies in the sub-region), which limited our ability to assess the representativeness of both these studies and our estimates for the broader sub-region.

Limitations related to study representativeness and reporting, and adjustments made in our analysis, may have influenced our estimates. Given the lack of population-representative studies, we used ANC attendees as the reference category for our prevalence estimates. At least 65% of studies focused exclusively on sexually active individuals, including antenatal, family planning, and gynaecology clinic attendees, and HIV/STI prevention trial participants. We considered these groups to be representative of the general population, which may result in overestimating STI prevalence compared to the total adult population. Variability in reporting across studies hindered our adjustment of estimates for HIV prevalence, granular age groups, or urban/rural location, which likely influenced sub-regional heterogeneities. Furthermore, our adjustments for diagnostic test performance may have overestimated prevalence, particularly for trichomoniasis, due to the poor sensitivity of widely used wet mount microscopy. Our reliance on previously published WHO performance data, without accounting for improvements in diagnostic accuracy over time, may have resulted in over-adjusting recent study observations and thus overestimating temporal trends.

Despite these limitations, our systematic review provides a comprehensive analysis of chlamydia, gonorrhoea, and trichomoniasis prevalence in SSA. We have addressed gaps in previous prevalence estimates by examining temporal trends and sub-regional variation, while incorporating studies from a range of populations considered representative of the general population. Our multiple analytical approaches yielded similar male-to-female prevalence ratio estimates, and our extensive sensitivity analyses of diagnostic test performance adjustments have strengthened the robustness of our findings.

In conclusion, we estimated a high and geographically varied prevalence of three curable STIs in SSA, with concerning increases in chlamydia prevalence over time. Male-to-female prevalence ratios for SSA and its sub-regions differed from existing WHO and GBD global ratios, emphasising the need to account for epidemiologic variations in future STI burden estimates and control strategies. Strengthening sex-stratified aetiologic surveillance, through both regular sentinel surveys and population-representative prevalence studies, is essential for progressing toward WHO targets and reducing the STI burden in SSA.

## Contributors

JM, JR, JWI-E, and M-CB conceptualised the study. DMR, EM, JM, JWI-E, JK, LH, MKW, OE, OS, QH, RLA, and SK screened abstracts and full text articles for inclusion. JM, LH, RLA, and QH extracted data from included studies. JM assessed diagnostic test validity, collated diagnostic test performance characteristics, and performed the data analysis. JM wrote the initial manuscript draft, which was revised with input from all authors. All authors approved the final manuscript.

## Data sharing

Data extracted from included studies and used for analysis are available as supplementary material. Code and data reproducing the analysis is available from https://github.com/juliamichalow/sti-prevalence-ratios.

## Data Availability

Source data were publicly available and submitted as a supplementary file.

