## Supplementary file for "Prevalence of chlamydia, gonorrhoea, and trichomoniasis among male and female general populations in sub-Saharan Africa from 2000-2024: A systematic review and meta-regression analysis"

### Supplementary material

Julia Michalow[1*], Lauren Hall[1], Jane Rowley PhD[2], Rebecca Anderson[1], Quinton Hayre[3], R Matthew Chico PhD[4], Olanrewaju Edun[1], Jesse Knight PhD[1], Salome Kuchukhidze PhD[3], Evidence Majaya[5], Domonique Reed PhD[3], Oliver Stevens[1], Magdelene K Walters[1], Remco PH Peters PhD[2], Anne Cori PhD[1], Prof Marie-Claude Boily PhD[1], Jeffrey W. Imai-Eaton PhD[1,3]

[1] MRC Centre for Global Infectious Disease Analysis, School of Public Health, Imperial College London, London, United Kingdom

[2] Department of Global HIV, Hepatitis and Sexually Transmitted Infections Programmes, World Health Organization, Geneva, Switzerland

[3] Center for Communicable Disease Dynamics, Department of Epidemiology, Harvard T.H. Chan School of Public Health, Boston, MA, USA

[4] Department of Disease Control, Faculty of Infectious and Tropical Diseases, London School of Hygiene & Tropical Medicine, London, United Kingdom

[5] Centre for Infectious Disease Epidemiology and Research, School of Public Health and Family Medicine, University of Cape Town, Cape Town, South Africa

S8 Adjusted prevalence ratios for chlamydia, gonorrhoea, and trichomoniasis in sub-Saharan Africa, estimated via log-binomial generalised linear mixed-effects models: *between-study sensitivity analysis using NAAT-diagnosed observations adjusted for test performance* . 21

S9 Adjusted prevalence ratios for chlamydia, gonorrhoea, and trichomoniasis in sub-Saharan Africa, estimated via log-binomial generalised linear mixed-effects models: *between-study sensitivity analysis using NAAT-diagnosed observations unadjusted for test performance* 22

S10 Adjusted prevalence ratios for chlamydia, gonorrhoea, and trichomoniasis in sub-Saharan Africa, estimated via log-binomial generalised linear mixed-effects models: *within-study sensitivity analysis using observations unadjusted for diagnostic test performance* . . . 23

S11 Adjusted prevalence ratios for chlamydia, gonorrhoea, and trichomoniasis in sub-Saharan Africa, estimated via log-binomial generalised linear mixed-effects models: *within-study sensitivity analysis using NAAT-diagnosed observations adjusted for test performance* . 24

S12 Adjusted prevalence ratios for chlamydia, gonorrhoea, and trichomoniasis in sub-Saharan Africa, estimated via log-binomial generalised linear mixed-effects models: *within-study sensitivity analysis using NAAT-diagnosed observations unadjusted for test performance* 25

**Table S1:** Search strategy for systematic review

**EMBASE:** Search conducted 17 September 2024 with 3794 articles retrieved.

**STI domain:** exp gonorrhea/ or exp Neisseria gonorrhoeae/ or exp Chlamydia trachomatis/ or exp Chlamydia trachomatis infection/ or exp vaginal trichomoniasis/ or exp trichomoniasis/ or exp Trichomonas vaginalis/ or (gonorrhea* or gonorrhoea* or gonorrhoeae* or chlamydia* or trichomonas or trichomoniasis).ab,ti,kw.

**AND sub-Saharan Africa domain:** exp Africa south of the Sahara/ or exp africa, eastern/ or exp africa, western/ or exp africa, central/ or exp africa, southern/ or (Africa or Angola or Benin or Botswana or Motswana or Batswana or Burkina or Burundi or "Cabo Verde" or "Cape verde" or Cameroon or "Central African Republic" or "Republique centrafricaine" or Chad or Comoros or Congo or "Democratic Republic of Congo" or "Republique democratique du Congo" or DRC or "Cote d'Ivoire" or "Ivory Coast" or Djibouti or Guinea or Eritrea or Erythree or Ethiopia or Gabon or Gambia or Ghana or Guinea or "Equatorial Guinea" or "Guinee Equatoriale" or "Equatoguinean" or "Guinea-Bissau" or Kenya or Lesotho or Basotho or Liberia or Madagascar or Malawi or Mali or Mauritania or Mozambique or Namibia or Niger or Nigeria or Rwanda or Rouanda or Ruanda or "Sao Tome" or Senegal or Seychelles or "Sierra Leone" or Somalia or Somali or "South Africa" or "Afrique du Sud" or "South Sudan" or "Soudan de sud" or Sudan or Swaziland or eSwatini or Tanzania or Togo or Uganda or Ouganda or Zambia or Zimbabwe).mp.

**AND** Limit publication year = "January 1, 2000 - Current"

**PubMed:** Search conducted 17 September 2024 with 2663 articles retrieved.

**STI domain:** "Neisseria gonorrhoeae"[MeSH Terms] OR "Gonorrhea"[MeSH Terms] OR "Chlamydia Infections"[MeSH Terms] OR "Chlamydia"[MeSH Terms] OR "Chlamydia trachomatis"[MeSH Terms] OR "Trichomonas vaginalis"[MeSH Terms] OR "Trichomonas Infections"[MeSH Terms] OR "Trichomonas"[MeSH Terms] OR "Trichomonas Vaginitis"[MeSH Terms] or gonorrhea*[Title/Abstract] OR gonorrhoea*[Title/Abstract] OR gonorrhoeae*[Title/Abstract] OR chlamydia*[Title/Abstract] OR trichomonas[Title/Abstract] OR trichomoniasis[Title/Abstract]

**AND sub-Saharan Africa domain:** "Africa south of the Sahara"[MeSH Terms] OR "africa, eastern"[MeSH Terms] OR "africa, western"[MeSH Terms] OR "africa, central"[MeSH Terms] OR "africa, southern"[MeSH Terms] OR "Africa"[All fields] OR "Angola"[All fields] OR "Benin"[All fields] OR "Botswana"[All fields] OR "Motswana"[All fields] OR "Batswana"[All fields] OR "Burkina"[All fields] OR "Burundi"[All fields] OR "Cabo Verde"[All fields] OR "Cape verde"[All fields] OR "Cameroon"[All fields] OR "Central African Republic"[All fields] OR "Republique centrafricaine"[All fields] OR "Chad"[All fields] OR "Comoros"[All fields] OR "Congo"[All fields] OR "Democratic Republic of Congo"[All fields] OR "Republique democratique du Congo"[All fields] OR "DRC"[All fields] OR "Cote d'Ivoire"[All fields] OR "Ivory Coast"[All fields] OR "Djibouti"[All fields] OR "Guinea"[All fields] OR "Eritrea"[All fields] OR "Ethiopia"[All fields] OR "Gabon"[All fields] OR "Gambia"[All fields] OR "Ghana"[All fields] OR "Guinea"[All fields] OR "Equatorial Guinea"[All fields] OR "Equatoguinean"[All fields] OR "Guinea-Bissau"[All fields] OR "Kenya"[All fields] OR "Lesotho"[All fields] OR "Basotho"[All fields] OR "Liberia"[All fields] OR "Madagascar"[All fields] OR "Malawi"[All fields] OR "Mali"[All fields] OR "Mauritania"[All fields] OR "Mozambique"[All fields] OR "Namibia"[All fields] OR "Niger"[All fields] OR "Nigeria"[All fields] OR "Rwanda"[All fields] OR "Rouanda"[All fields] OR "Ruanda"[All fields] OR "Sao Tome"[All fields] OR "Senegal"[All fields] OR "Seychelles"[All fields] OR "Sierra Leone"[All fields] OR "Somalia"[All fields] OR "Somali"[All fields] OR "South Africa"[All fields] OR "Afrique du Sud"[All fields] OR "South Sudan"[All fields] OR "Sudan"[All fields] OR "Swaziland"[All fields] OR "eSwatini"[All fields] OR "Tanzania"[All fields] OR "Togo"[All fields] OR "Uganda"[All fields] OR "Ouganda"[All fields] OR "Zambia"[All fields] OR "Zimbabwe"[All fields]

**AND** Limit publication year = "January 1, 2000 - Current"

**Global Health:** Search conducted 17 September 2024 with 2364 articles retrieved.

**STI domain:** exp gonorrhoea/ or exp Neisseria gonorrhoeae/ or exp Chlamydia trachomatis/ or exp Chlamydia/ or exp trichomoniasis/ or exp Trichomonas vaginalis/ or (gonorrhea* or gonorrhoea* or gonorrhoeae* or chlamydia* or trichomonas or trichomoniasis).ab,ti,mp.

**AND sub-Saharan Africa domain:** exp ”Africa South of Sahara”/ or exp East Africa/ or exp Africa/ or exp Central Africa/ or exp Southern Africa/ or exp West Africa/ or (Africa or Angola or Benin or Botswana or Motswana or Batswana or Burkina or Burundi or ”Cabo Verde” or ”Cape verde” or Cameroon or ”Central African Republic” or ”Republique centrafricaine” or Chad or Comoros or Congo or ”Democratic Republic of Congo” or ”Republique democratique du Congo” or DRC or ”Cote d’Ivoire” or ”Ivory Coast” or Djibouti or Guinea or Eritrea or Erythree or Ethiopia or Gabon or Gambia or Ghana or Guinea or ”Equatorial Guinea” or ”Guinee Equatoriale” or ”Equatoguinean” or ”Guinea-Bissau” or Kenya or Lesotho or Basotho or Liberia or Madagascar or Malawi or Mali or Mauritania or Mozambique or Namibia or Niger or Nigeria or Rwanda or Rouanda or Ruanda or ”Sao Tome” or Senegal or Seychelles or ”Sierra Leone” or Somalia or Somali or ”South Africa” or ”Afrique du Sud” or ”South Sudan” or ”Soudan de sud” or Sudan or Swaziland or eSwatini or Tanzania or Togo or Uganda or Ouganda or Zambia or Zimbabwe).mp.

**AND** Limit publication year = ”January 1, 2000 - Current”

**MEDLINE:** Search conducted 17 September 2024 with 2231 articles retrieved.

**STI domain:** exp Neisseria gonorrhoeae/ or exp Gonorrhea/ or exp Chlamydia Infections/ or exp Chlamydia/ or exp Chlamydia trachomatis/ or exp Trichomonas vaginalis/ or exp Trichomonas Infections/ or exp Trichomonas/ or exp Trichomonas Vaginitis/ or (gonorrhea* or gonorrhoea* or gonorrhoeae* or chlamydia* or trichomonas* or trichomoniasis*).ab,ti,kw.

**AND sub-Saharan Africa domain:** exp Africa, Western/ or exp Africa, Central/ or exp ”Africa South of the Sahara”/ or exp Africa/ or exp Africa, Eastern/ or exp Africa, Southern/ or (Africa or Angola or Benin or Botswana or Motswana or Batswana or Burkina or Burundi or ”Cabo Verde” or ”Cape verde” or Cameroon or ”Central African Republic” or ”Republique centrafricaine” or Chad or Comoros or Congo or ”Democratic Republic of Congo” or ”Republique democratique du Congo” or DRC or ”Cote d’Ivoire” or ”Ivory Coast” or Djibouti or Guinea or Eritrea or Erythree or Ethiopia or Gabon or Gambia or Ghana or Guinea or ”Equatorial Guinea” or ”Guinee Equatoriale” or ”Equatoguinean” or ”Guinea-Bissau” or Kenya or Lesotho or Basotho or Liberia or Madagascar or Malawi or Mali or Mauritania or Mozambique or Namibia or Niger or Nigeria or Rwanda or Rouanda or Ruanda or ”Sao Tome” or Senegal or Seychelles or ”Sierra Leone” or Somalia or Somali or ”South Africa” or ”Afrique du Sud” or ”South Sudan” or ”Soudan de sud” or Sudan or Swaziland or eSwatini or Tanzania or Togo or Uganda or Ouganda or Zambia or Zimbabwe).mp.

**AND** Limit publication year = ”January 1, 2000 - Current”

**African Index Medicus:** Search conducted 17 September 2024 with 84 articles retrieved.

**STI domain:** mh:(”Chlamydia Infections” OR ”Chlamydia Infection” OR ”Chlamydia trachomatis” OR “Chlamydia”, OR ”Gonorrhoea” OR “Neisseria gonorrhoeae” OR “Trichomonas” OR “Trichomoniasis” OR “Trichomonas vaginalis” OR “Trichomonas infections” OR “Trichomonas infection”) or tw:(gonorrhea* or gonorrhoea* or gonorrhoeae* or chlamydia* or trichomonas or trichomoniasis)

**AND** Limit publication year = ”January 1, 2000 - Current”

**Table S2:** Classification of included countries by sub-region in sub-Saharan Africa

| Sub-region | Country |
|---|---|
| Central Africa | Angola, Cameroon, Central African Republic, Chad, Congo, Democratic Republic of Congo, Equatorial Guinea, Gabon, Sao Tome and Principe |
| Western Africa | Benin, Burkina Faso, Cabo Verde, Cote d'Ivoire, Gambia, Ghana, Guinea-Bissau, Guinea, Liberia, Mali, Mauritania, Niger, Nigeria, Senegal, Sierra Leone, Togo |
| Eastern Africa | Burundi, Comoros, Djibouti, Eritrea, Ethiopia, Kenya, Madagascar, Malawi, Mauritius, Mozambique, Rwanda, Seychelles, Somalia, South Sudan, Uganda, United Republic of Tanzania, Zambia, Zimbabwe |
| Southern Africa | Botswana, Eswatini, Lesotho, Namibia, South Africa |

Countries classified according to UN M49 Standard.[1]

**Table S3:** Variables extracted from included studies

| Category | Variable |
|---|---|
| Study characteristics | • Authors<br>• Publication title<br>• Publication year<br>• Dates of data collection<br>• Country of study<br>• Sub-national region or city of study<br>• Study name (as applicable) |
| Participant characteristics | • Study population category<br>• Study population age (mean, SD, median, IQR, range)<br>• Study population HIV status<br>• Study population HIV prevalence |
| Diagnostic methodology | • Diagnostic test per infection<br>• Diagnostic specimen per infection<br>• Number tested per infection<br>• Number positive per infection<br>• Prevalence of each infection |

**Table S4:** Diagnostic test performance characteristics

| STI | Test | Specimen | N | Sensitivity (%) | Specificity (%) | Source |
|---|---|---|---|---|---|---|
| **Female** | | | | | | |
| CT | DFA | Genital fluid | 2 | 82.0 | 98.5 | WHO 1999[2] |
| | DFA | Urine | 1 | 82.0 | 98.5 | WHO 1999[2] |
| | ELISA | Genital fluid | 2 | 65.0 | 100.0 | WHO 2011[3] |
| | NAAT | Genital fluid | 105 | 88.6 | 99.6 | WHO 2011[3] |
| | NAAT | Urine | 41 | 87.0 | 99.8 | WHO 2011[3] |
| | NAAT | Genital fluid or urine | 9 | 87.0 | 99.6 | Combined estimate[3,i] |
| | Rapid antigen test | Genital fluid | 8 | 56.0 | 99.0 | Grillo-Ardila 2020,[4] Zhou 2021[5] |
| | DFA and NAAT | genital fluid | 1 | 82.0 | 99.6 | Combined estimate[2,3,ii] |
| NG | Culture | Genital fluid | 23 | 75.7 | 100.0 | WHO 2011[3] |
| | NAAT | Genital fluid | 98 | 93.3 | 99.2 | WHO 2011[3] |
| | NAAT | Urine | 40 | 91.6 | 100.0 | WHO 2011[3] |
| | NAAT | Genital fluid or urine | 8 | 91.6 | 99.2 | Combined estimate[3,i] |
| | Rapid antigen test | Genital fluid | 1 | 70.0 | 96.0 | Watchirs Smith 2013[6] |
| | Culture or NAAT | Genital fluid or urine | 1 | 75.7 | 99.2 | Combined estimate[3,i] |
| TV | Culture | Genital fluid | 32 | 68.8 | 100.0 | WHO 2011[3] |
| | Culture | Urine | 1 | 68.8 | 100.0 | Assumed equivalent to genital fluid test. |
| | NAAT | Genital fluid | 47 | 95.0 | 98.0 | WHO 2011[3] |
| | NAAT | Urine | 7 | 66.9 | 98.3 | WHO 2011[3] |
| | NAAT | Genital fluid or urine | 1 | 66.9 | 98.0 | Combined estimate[3,i] |
| | Rapid antigen test | Genital fluid | 17 | 83.3 | 98.8 | Gaydos 2017[7] |
| | Wet mount | Genital fluid | 71 | 52.0 | 100.0 | WHO 2011[3] |
| | Wet mount | Urine | 2 | 52.0 | 100.0 | Assumed equivalent to genital fluid test. |
| | Wet mount | Genital fluid and urine | 4 | 52.0 | 100.0 | Assumed equivalent to genital fluid test. |
| | Culture and wet mount | Genital fluid | 5 | 52.0 | 100.0 | Combined estimate[3,ii] |
| | Culture or NAAT | Genital fluid or urine | 1 | 66.9 | 98.0 | Combined estimate[3,i] |
| | Wet mount and NAAT | Genital fluid | 1 | 52.0 | 100.0 | Combined estimate[3,ii] |
| **Male** | | | | | | |
| CT | NAAT | Genital fluid | 1 | 87.5 | 99.2 | WHO 2011[3] |
| | NAAT | Urine | 32 | 87.8 | 99.3 | WHO 2011[3] |
| | DFA and NAAT | Genital fluid | 1 | 82.0 | 99.2 | Combined estimate[2,3,ii] |
| NG | Culture | Genital fluid | 1 | 87.6 | 100.0 | WHO 2011[3] |
| | NAAT | Genital fluid | 1 | 96.1 | 99.0 | WHO 2011[3] |
| | NAAT | Urine | 29 | 80.9 | 99.9 | WHO 2011[3] |
| | Culture or NAAT | Urine | 1 | 80.9 | 99.9 | Combined estimate[3,i] |
| TV | Culture | Urine | 3 | 87.6 | 100.0 | WHO 2011[3] |
| | Culture | Genital fluid or urine | 1 | 87.6 | 100.0 | Combined estimate[3,i] |
| | NAAT | Urine | 13 | 96.0 | 97.7 | WHO 2011[3] |
| | Rapid antigen test | Genital fluid | 1 | 68.5 | 97.4 | Assumed equivalent to test for females. |
| | Wet mount | Urine | 1 | 44.0 | 100.0 | WHO 2011[3] |
| | Culture or NAAT | Urine | 1 | 87.6 | 97.7 | Combined estimate[3,i] |

N: Number of observations. CT: *Chlamydia trachomatis*, NG: *Neisseria gonorrhoeae*, TV: *Trichomonas vaginalis.* DFA: Direct fluorescent antibody, ELISA: Enzyme-linked immunosorbent assay, NAAT: Nucleic acid amplification test.
Sensitivity and specificity values collated per approach in Michalow 2024.[8] Combined performance characteristics were estimated by: (i) using the lower sensitivity and lower specificity values when at least one of two diagnostic approaches identified a positive case, and (ii) using the lower sensitivity and higher specificity values when both diagnostic approaches needed to be positive.

**Table S5:** Overview of studies included in the systematic review

| Region | Reference | Year | Country | Population | Sex | Age | Age range | HIV status | Infections |
|---|---|---|---|---|---|---|---|---|---|
| WA | Lagarde 2004[9] | 2000 | Burkina Faso | Population-representative survey participants | F, M | Adult | 13 to 49 | Non-stratified | CT, NG, TV |
| WA | Lafort 2003[10] | 2000 | Cote d'Ivoire | FP attendees | F | Adult | 18 to 53 | Non-stratified | CT, NG, TV |
| WA | Aboyeji 2003[11] | 2000 | Nigeria | ANC attendees | F | Adult | 19 to 43 | Non-stratified | NG, TV |
| WA | Donbraye 2010[12] | 2000 | Nigeria | ANC attendees | F | Adult | NR | Non-stratified | TV |
| WA | Apea-Kubi 2004[13] | 2001 | Ghana | ANC attendees | F | Adult | 16+ | Non-stratified | CT, NG |
| WA | Apea-Kubi 2005[14] | 2002 | Ghana | ANC attendees | F | Adult | 16+ | Non-stratified | TV |
| WA | Adejuwon 2005[15] | 2002* | Nigeria | FP attendees | F | Adult | NR | Non-stratified | NG, TV |
| WA | Obiajuru 2005[16] | 2002 | Nigeria | Community members | F | Adult | NR | Non-stratified | NG, TV |
| WA | Tukur 2006[17] | 2002 | Nigeria | FP attendees | F | Adult | NR | Non-stratified | CT |
| WA | Balaka 2005[18] | 2002 | Togo | ANC attendees | F | Adult | 16 to 42 | Non-stratified | TV |
| WA | Kirakoya-Samadoulougou 2008[19] | 2003 | Burkina Faso | ANC attendees | F | Adult | 14 to 49 | Non-stratified | TV |
| WA | Siemer 2008[20] | 2003 | Ghana | ANC attendees | F | Adult | NR | Non-stratified | CT |
| WA | Chigbu 2006[21] | 2003* | Nigeria | ANC attendees, GYN attendees | F | Adult | 15 to 65 | Non-stratified | NG, TV |
| WA | Inabo 2006[22] | 2003* | Nigeria | ANC attendees | F | Adult | 18 to 47 | Non-stratified | TV |
| WA | Jatau 2006[23] | 2003* | Nigeria | ANC attendees | F | Adult | 16+ | Non-stratified | TV |
| WA | Sagay 2005[24] | 2003 | Nigeria | ANC attendees | F | Adult | NR | Non-stratified | TV |
| WA | Yirenya-Tawiah 2014[25] | 2006 | Ghana | Community members | F | Adult | 15 to 49 | Non-stratified | CT, NG |
| WA | Fayemiwo 2018[26] | 2006 | Nigeria | FP attendees | F | Adult | 19 to 54 | Non-stratified | CT, NG, TV |
| WA | Omoregie 2009[27] | 2006* | Nigeria | PHC/OPD attendees | F, M | Adult | NR | HIV negative | TV |
| WA | Kengne 2010[28] | 2007* | Cote d'Ivoire | ANC attendees | F | Adult | NR | Non-stratified | CT, NG, TV |
| WA | Chinyere 2012[29] | 2007 | Nigeria | ANC attendees | F | Adult | 15 to 40 | Non-stratified | TV |
| WA | Niemogha 2010[30] | 2007* | Nigeria | FP attendees, GYN attendees, Students | F | Adult | NR | Non-stratified | TV |

*Continued...*

| Region | Reference | Year | Country | Population | Sex | Age | Age range | HIV status | Infections |
|---|---|---|---|---|---|---|---|---|---|
| WA | Behanzin 2012[31] | 2008 | Benin | Population-representative survey participants | F, M | Adult | 15 to 49 | Non-stratified | CT, NG |
| WA | Usanga 2011[32] | 2008 | Nigeria | ANC attendees | F | Adult | 15 to 49 | Non-stratified | NG, TV |
| WA | Sam-Wobo 2012[33] | 2009* | Nigeria | ANC attendees | F | Adult | 16 to 50 | Non-stratified | TV |
| WA | Arinze 2014[34] | 2011* | Nigeria | Students | F | Adult | 15 to 30 | Non-stratified | CT |
| WA | Bolaji 2013[35] | 2011 | Nigeria | ANC attendees | F | Adult | 20 to 40 | Non-stratified | TV |
| WA | Tchelougou 2013[36] | 2011 | Togo | ANC attendees | F | Adult | NR | Non-stratified | TV |
| WA | Volker 2017[37] | 2012 | Ghana | ANC attendees | F | Adult | 14 to 48 | Non-stratified | CT, NG |
| WA | Adesiji 2015[38] | 2012* | Nigeria | FP attendees | F | Adult | 20+ | Non-stratified | CT |
| WA | Olowe 2014[39] | 2012 | Nigeria | ANC attendees | F | Adult | 21 to 40 | Non-stratified | TV |
| WA | Samuel 2015[40] | 2012* | Nigeria | ANC attendees | F | Adult | 21 to 50 | Non-stratified | TV |
| WA | Etuketu 2015[41] | 2013 | Nigeria | ANC attendees | F | Adult | 15 to 44 | Non-stratified | TV |
| WA | Nnaemeka 2016[42] | 2013* | Nigeria | Population-representative survey participants | F | Adult | 22 to 42 | Non-stratified | TV |
| WA | Olusegun-Joseph 2016[43] | 2013* | Nigeria | PHC/OPD attendees, Students | F | Adult | 16 to 55 | Non-stratified | TV |
| WA | Akinbo 2017[44] | 2014* | Nigeria | Students | F | Youth | 13 to 17 | Non-stratified | TV |
| WA | Oyeyemi 2016[45] | 2014 | Nigeria | ANC attendees | F | Adult | 21+ | Non-stratified | TV |
| WA | Wokem 2015[46] | 2014 | Nigeria | ANC attendees | F | Adult | 11 to 60 | Non-stratified | TV |
| WA | Sangare 2021[47] | 2015 | Burkina Faso | ANC attendees | F | Adult | 15 to 49 | Non-stratified | TV |
| WA | Konadu 2019[48] | 2015 | Ghana | ANC attendees | F | Adult | 12 to 54 | Non-stratified | TV |
| WA | Alexander 2018[49] | 2015 | Nigeria | ANC attendees | F | Adult | 15 to 60 | Non-stratified | TV |
| WA | Ebhodaghe 2017[50] | 2015 | Nigeria | ANC attendees | F | Adult | 19 to 43 | HIV negative | CT, NG, TV |
| WA | Asmah 2017[51] | 2016 | Ghana | ANC attendees | F | Adult | NR | Non-stratified | TV |
| WA | Squire 2019[52] | 2016 | Ghana | GYN attendees | F | Adult | 16+ | Non-stratified | TV |
| WA | Cowley 2021[53] | 2016 | Guinea-Bissau | Population-representative survey participants | F, M | Adult | 16 to 49 | Non-stratified | CT, NG, TV |
| WA | Ezeanya 2019[54] | 2016* | Nigeria | Students | F | Adult | 15 to 39 | Non-stratified | CT, TV |
| WA | Ukatu 2019[55] | 2016* | Nigeria | ANC attendees | F | Adult | 18 to 45 | Non-stratified | TV |

*Continued...*

| Region | Reference | Year | Country | Population | Sex | Age | Age range | HIV status | Infections |
|---|---|---|---|---|---|---|---|---|---|
| WA | Kashibu 2018[56] | 2017 | Nigeria | ANC attendees | F | Adult | 15 to 39 | Non-stratified | TV |
| WA | Odaranle 2020[57] | 2017 | Nigeria | FP attendees | F | Adult | 20 to 45 | Non-stratified | TV |
| WA | Isara 2021[58] | 2017 | The Gambia | ANC attendees | F | Adult | 15 to 44 | Non-stratified | CT, NG, TV |
| WA | Jary 2021[59] | 2018 | Mali | PHC/OPD attendees | F | Adult | 18+ | Non-stratified | CT, NG, TV |
| WA | Rasheed 2021[60] | 2018 | Nigeria | ANC attendees | F | Adult | 18+ | Non-stratified | TV |
| WA | Ajani 2022[61] | 2019 | Nigeria | Students | F, M | Adult | 15 to 30 | Non-stratified | TV |
| WA | Auta 2020[62] | 2019 | Nigeria | ANC attendees | F | Adult | 15+ | Non-stratified | TV |
| WA | Maureen 2022[63] | 2019* | Nigeria | ANC attendees | F | Adult | 18+ | Non-stratified | CT, NG, TV |
| WA | Lingani 2021[64] | 2020 | Burkina Faso | ANC attendees | F | Adult | 16 to 45 | Non-stratified | CT |
| WA | Agabi 2023[65] | 2020* | Nigeria | GYN attendees | F | Adult | 16 to 57 | Non-stratified | TV |
| WA | Enwuru 2024[66] | 2020 | Nigeria | ANC attendees | F | Adult | 15+ | HIV negative | TV |
| WA | Butcher 2023[67] | 2020 | The Gambia | Prevention trial participants | F | Adult | 15 to 69 | Non-stratified | CT, NG, TV |
| WA | Ngom 2023[68] | 2021 | Senegal | ANC attendees | F | Adult | 16 to 46 | Non-stratified | NG, TV |
| CA | Ngandjio 2003[69] | 2001 | Cameroon | Students | F, M | Youth | NR | Non-stratified | CT |
| CA | Kinoshita-Moleka 2008[70] | 2004 | Democratic Republic of Congo | ANC attendees | F | Adult | 15 to 45 | Non-stratified | CT, NG |
| CA | Mbu 2008[71] | 2006 | Cameroon | ANC attendees | F | Adult | NR | Non-stratified | CT, NG, TV |
| CA | Alexandre 2015[72] | 2012 | Angola | PHC/OPD attendees | F | Adult | 14 to 52 | Non-stratified | CT, NG |
| CA | Vieira-Baptista 2017[73] | 2015 | Sao Tome and Principe | GYN attendees | F | Adult | 21 to 60 | Non-stratified | CT, NG, TV |
| CA | Compain 2019[74] | 2017 | Chad | Community members | F | Adult | 20 to 65 | Non-stratified | CT, NG, TV |
| CA | Nodjikouambaye 2019[75] | 2017 | Chad | GYN attendees | F | Adult | 18+ | Non-stratified | CT, NG, TV |
| CA | Gadoth 2019[76] | 2017 | Democratic Republic of Congo | ANC attendees | F | Adult | 18+ | HIV negative | CT, NG, TV |
| CA | Mbah 2022[77] | 2018 | Cameroon | ANC attendees | F | Adult | 15 to 46 | Non-stratified | CT, NG, TV |
| CA | Payne 2020[78] | 2018 | Cameroon | PHC/OPD attendees | F | Adult | 15 to 55 | Non-stratified | TV |
| CA | Ngombe Mouabata 2024[79] | 2021 | Congo | GYN attendees | F | Adult | 21 to 71 | Non-stratified | CT |
| CA | Eyong 2023[80] | 2022 | Cameroon | PHC/OPD attendees | F | Adult | 17 to 53 | Non-stratified | TV |

*Continued...*

| Region | Reference | Year | Country | Population | Sex | Age | Age range | HIV status | Infections |
|---|---|---|---|---|---|---|---|---|---|
| EA | Hawken 2002[81] | 2000 | Kenya | Population-representative survey participants | F, M | Adult | 15 to 49 | Non-stratified | CT, NG |
| EA | Kaydos-Daniels 2003[82] | 2000 | Malawi | PHC/OPD attendees | M | Adult | 18+ | Non-stratified | TV |
| EA | Paz-Soldan 2012[83] | 2000 | Malawi | Population-representative survey participants | F, M | Adult | 15-44 | Non-stratified | CT, NG |
| EA | Menendez 2010[84] | 2000 | Mozambique | ANC attendees | F | Adult | 14 to 61 | Non-stratified | CT, NG, TV |
| EA | Clift 2003[85] | 2000 | Tanzania | Community members | F, M | Adult | 16 to 54 | Non-stratified | CT, NG |
| EA | Cowan 2002[86] | 2000 | Zimbabwe | Community members | F, M | Youth | 16 to 19 | Non-stratified | CT, NG |
| EA | van de Wijgert 2009[87] | 2002 | Uganda, Zimbabwe | FP attendees | F | Adult | 18 to 35 | HIV negative | CT, NG, TV |
| EA | Munjoma 2010[88] | 2002 | Zimbabwe | ANC attendees | F | Adult | NR | Non-stratified | TV |
| EA | Bailey 2007[89] | 2003 | Kenya | Prevention trial participants | M | Youth | 18 to 24 | HIV negative | CT, NG, TV |
| EA | Ghebremichael 2009[90] | 2003 | Tanzania | Community members | F | Adult | 20 to 44 | Non-stratified | CT, NG, TV |
| EA | Ghebremichael 2011[91] | 2003 | Tanzania | Community members | M | Adult | 20+ | Non-stratified | CT, TV |
| EA | Mapingure 2010[92] | 2003 | Tanzania, Zimbabwe | ANC attendees | F | Adult | 14 to 43 | Non-stratified | TV |
| EA | Msuya 2009[93] | 2003 | Tanzania | ANC attendees | F | Adult | 14 to 43 | Non-stratified | NG, TV |
| EA | Celentano 2010[94] | 2003 | Zimbabwe | Community members | F, M | Adult | 18 to 30 | Non-stratified | CT, NG, TV |
| EA | Mensch 2008[95] | 2004 | Malawi | Population-representative survey participants | F | Youth | 15 to 21 | Non-stratified | CT, NG, TV |
| EA | Lujan 2008[96] | 2004 | Mozambique | ANC attendees | F | Adult | 15 to 45 | Non-stratified | CT, NG |
| EA | Kamali 2010[97] | 2004 | Uganda | Prevention trial participants | F | Adult | 18 to 45 | Non-stratified | CT, NG, TV |
| EA | Tann 2006[98] | 2004 | Uganda | ANC attendees | F | Adult | 15 to 40 | Non-stratified | CT, NG, TV |
| EA | Ramjee 2008[99] | 2004 | Zambia | Prevention trial participants | F | Adult | 18+ | Non-stratified | CT, NG, TV |
| EA | Venkatesh 2011[100] | 2004 | Zimbabwe | Prevention trial participants | F | Adult | 18 to 49 | HIV negative | CT, NG, TV |
| EA | Gray 2009[101] | 2005 | Uganda | Community members | F | Adult | 15 to 49 | HIV negative | TV |

*Continued...*

| Region | Reference | Year | Country | Population | Sex | Age | Age range | HIV status | Infections |
|---|---|---|---|---|---|---|---|---|---|
| EA | Lingappa 2009[102] | 2006 | Botswana, Kenya, Rwanda, South Africa, Tanzania, Uganda, Zambia | Prevention trial participants | F, M | Adult | 18+ | HIV negative | CT, NG, TV |
| EA | Chersich 2009[103] | 2006 | Kenya | PHC/OPD attendees | F | Adult | 16 to 45 | Non-stratified | TV |
| EA | Guffey 2014[104] | 2006 | Malawi, Zambia, Zimbabwe | Prevention trial participants | F | Adult | 18+ | HIV negative | CT, NG, TV |
| EA | McCormack 2010[105] | 2007 | Tanzania, Uganda, Zambia | Prevention trial participants | F | Adult | 16+ | HIV negative | CT, NG, TV |
| EA | Crucitti 2010[106] | 2007* | Zambia | ANC attendees, Students | F | Adult | 15 to 42 | Non-stratified | TV |
| EA | Otieno 2015[107] | 2008 | Kenya | Community members | F, M | Adult | 18 to 34 | HIV negative | CT, NG |
| EA | Mocumbi 2017[108] | 2008 | Mozambique | Prevention trial participants | F | Adult | 17 to 59 | HIV negative | CT, NG, TV |
| EA | Lemme 2013[109] | 2008 | Tanzania | Community members | F, M | Youth | 15 to 30 | Non-stratified | CT, NG |
| EA | Muvunyi 2011[110] | 2009 | Rwanda | Community members | F | Adult | 21 to 45 | Non-stratified | CT, NG |
| EA | Rutherford 2014[111] | 2009 | Uganda | Students | F, M | Youth | 19 to 25 | Non-stratified | CT, NG, TV |
| EA | Ademe 2013[112] | 2010 | Ethiopia | ANC attendees | F | Adult | 15 to 49 | Non-stratified | TV |
| EA | Chiduo 2012[113] | 2010 | Tanzania | ANC attendees | F | Adult | 18 to 44 | Non-stratified | CT, NG, TV |
| EA | Downs 2012[114] | 2010 | Tanzania | PHC/OPD attendees | F | Adult | 18 to 50 | Non-stratified | CT, NG |
| EA | Jespers 2014[115] | 2011 | Kenya | ANC attendees, FP attendees | F | Adult | 18 to 35 | Non-stratified | CT, NG, TV |
| EA | de Walque 2012[116] | 2011 | Tanzania | Prevention trial participants | F, M | Adult | 18 to 30 | Non-stratified | CT, NG, TV |
| EA | Lazenby 2014[117] | 2011* | Tanzania | GYN attendees | F | Adult | 30 to 60 | Non-stratified | CT, NG, TV |
| EA | Kiene 2017[118] | 2011 | Uganda | PHC/OPD attendees | F, M | Adult | 18+ | Non-stratified | CT, NG |
| EA | Ogilvie 2013[119] | 2011 | Uganda | Community members | F | Adult | 26 to 69 | Non-stratified | CT, NG |
| EA | Eshete 2013[120] | 2012 | Ethiopia | ANC attendees | F | Adult | 15 to 36 | Non-stratified | TV |
| EA | Kerubo 2016[121] | 2012 | Kenya | Students | F | Youth | 14 to 17 | Non-stratified | CT, NG, TV |
| EA | Kinuthia 2015[122] | 2012 | Kenya | ANC attendees | F | Adult | 14+ | HIV negative | CT, NG, TV |
| EA | Ravindran 2021[123] | 2012 | Kenya | ANC attendees | F | Adult | 14+ | HIV negative | CT, NG, TV |
| EA | Nkhoma 2017[124] | 2012 | Malawi | ANC attendees | F | Adult | 15+ | Non-stratified | TV |

*Continued...*

| Region | Reference | Year | Country | Population | Sex | Age | Age range | HIV status | Infections |
|---|---|---|---|---|---|---|---|---|---|
| EA | Hokororo 2015[125] | 2012 | Tanzania | ANC attendees | F | Youth | 14 to 20 | Non-stratified | CT, NG, TV |
| EA | Stephen 2017[126] | 2012 | Zimbabwe | ANC attendees | F | Adult | 18+ | Non-stratified | CT |
| EA | Mulu 2015[127] | 2013 | Ethiopia | ANC attendees | F | Adult | 15 to 49 | Non-stratified | NG, TV |
| EA | Maina 2016[128] | 2013 | Kenya | FP attendees | F | Adult | 20 to 49 | Non-stratified | CT, NG, TV |
| EA | Palanee-Phillips 2015[129] | 2013 | Malawi, Uganda, Zimbabwe | Prevention trial participants | F | Adult | 18 to 45 | HIV negative | CT, NG, TV |
| EA | Kestelyn 2018[130] | 2013 | Rwanda | Prevention trial participants | F | Adult | 18 to 35 | Non-stratified | CT, NG, TV |
| EA | Donders 2016[131] | 2013* | Uganda | PHC/OPD attendees | F | Adult | NR | Non-stratified | CT, NG, TV |
| EA | Nakubulwa 2015[132] | 2013 | Uganda | ANC attendees | F | Adult | 18+ | Non-stratified | CT, TV |
| EA | Schonfeld 2018 | 2014 | Ethiopia | ANC attendees | F | Adult | NR | Non-stratified | CT, NG, TV |
| EA | Kanyina 2017[133] | 2014 | Kenya | GYN attendees | F | Adult | 15+ | Non-stratified | TV |
| EA | Oliver 2018[134] | 2014 | Kenya | Community members | F | Adult | 18 to 34 | Non-stratified | CT, NG |
| EA | Franceschi 2016[135] | 2014 | Rwanda | Students | F | Youth | 18 to 20 | Non-stratified | CT |
| EA | Francis 2019[136] | 2014 | Tanzania | Students | F | Youth | 17 to 18 | Non-stratified | CT, NG, TV |
| EA | Homsy 2019[137] | 2014 | Uganda | ANC attendees | F | Adult | 18 to 49 | HIV negative | TV |
| EA | Moses 2015[138] | 2014 | Uganda | Community members | F | Adult | 30 to 65 | Non-stratified | CT, NG |
| EA | Chaponda 2021[139] | 2014 | Zambia | ANC attendees | F | Adult | NR | Non-stratified | CT, NG, TV |
| EA | Tadesse 2016[140] | 2015 | Ethiopia | GYN attendees | F | Adult | 15 to 49 | Non-stratified | NG |
| EA | Masese 2017[141] | 2015 | Kenya | Students | F | Youth | 15 to 24 | Non-stratified | CT, NG, TV |
| EA | Masha 2017[142] | 2015 | Kenya | ANC attendees | F | Adult | 18 to 45 | Non-stratified | CT, NG, TV |
| EA | Yuh 2020[143] | 2015 | Kenya | Students | F | Youth | 16 to 20 | HIV negative | CT, NG, TV |
| EA | Mukanyangezi 2018[144] | 2015 | Rwanda | GYN attendees | F | Adult | 18+ | HIV negative | TV |
| EA | Maufi 2016[145] | 2015 | Tanzania | ANC attendees | F | Adult | 17 to 46 | Non-stratified | TV |
| EA | Yegorov 2018[146] | 2015 | Uganda | PHC/OPD attendees | F | Adult | 18 to 45 | HIV negative | CT, NG, TV |
| EA | Deese 2021[147] | 2016 | Kenya, Zambia | FP attendees | F | Adult | 16 to 35 | HIV negative | CT, NG |
| EA | Mgodi 2021[148] | 2017 | Kenya, Malawi, Mozambique, Tanzania, Zimbabwe | Prevention trial participants | F | Adult | 18 to 50 | HIV negative | CT, NG |

*Continued...*

| Region | Reference | Year | Country | Population | Sex | Age | Age range | HIV status | Infections |
|---|---|---|---|---|---|---|---|---|---|
| EA | Baussano 2021[149] | 2017 | Rwanda | Students | F | Youth | 17 to 21 | Non-stratified | CT |
| EA | Nsereko 2020[150] | 2017 | Rwanda | ANC attendees | F | Adult | 18 to 49 | Non-stratified | TV |
| EA | Masatu 2022[151] | 2017 | Tanzania | FP attendees | F | Adult | 18+ | Non-stratified | CT |
| EA | Kahsay 2023[152] | 2018 | Ethiopia | PHC/OPD attendees | F, M | Adult | 15+ | Non-stratified | NG |
| EA | Madanitsa 2023[153] | 2018 | Kenya, Malawi, Tanzania | ANC attendees | F | Adult | NR | HIV negative | CT, NG, TV |
| EA | Mehta 2023[154] | 2018 | Kenya | Students | F | Youth | 14 to 22 | Non-stratified | CT, NG, TV |
| EA | Celum 2022[155] | 2019 | Kenya | FP attendees | F | Youth | 16 to 25 | HIV negative | CT, NG |
| EA | Lokken 2022[156] | 2019 | Kenya | FP attendees | F | Adult | 18 to 45 | HIV negative | CT, NG, TV |
| EA | Juliana 2020[157] | 2019 | Tanzania | ANC attendees | F | Adult | 16 to 48 | Non-stratified | CT, NG, TV |
| EA | Chitneni 2020[158] | 2019 | Uganda | PHC/OPD attendees | F | Adult | 18 to 40 | HIV negative | CT, NG, TV |
| EA | Grabowski 2022[159] | 2019 | Uganda | Population-representative survey participants | F, M | Adult | 18 to 49 | Non-stratified | CT, NG, TV |
| EA | Husen 2023[160] | 2020* | Ethiopia | ANC attendees | F | Adult | 17 to 37 | Non-stratified | TV |
| EA | Zenebe 2021[161] | 2020 | Ethiopia | ANC attendees | F | Adult | 17 to 41 | Non-stratified | CT, NG, TV |
| EA | Heffron 2021[162] | 2020 | Kenya | GYN attendees | F | Adult | 15 to 30 | HIV negative | CT, NG |
| EA | Mcharo 2022[163] | 2020 | Tanzania | Students | F, M | Youth | 18 to 24 | Non-stratified | CT, NG |
| EA | Nair 2023[164] | 2020 | Uganda, Zimbabwe | Prevention trial participants | F | Youth | 16 to 21 | HIV negative | CT, NG, TV |
| EA | Martin 2021[165] | 2020 | Zimbabwe | Community members | F, M | Youth | 16 to 24 | Non-stratified | CT, NG |
| EA | Nyakambi 2022[166] | 2021 | Kenya | PHC/OPD attendees | F | Adult | 18 to 49 | Non-stratified | CT |
| EA | Oware 2023[167] | 2021 | Kenya | Prevention trial participants | F | Adult | 18 to 30 | HIV negative | CT, NG |
| EA | van der Veer 2024[168] | 2021 | Malawi | ANC attendees | F | Adult | NR | Non-stratified | CT, NG, TV |
| EA | Sineque 2024[169] | 2021 | Mozambique | PHC/OPD attendees | F | Adult | 30 to 55 | Non-stratified | CT, NG |
| EA | Mbuvi 2024[170] | 2022 | Kenya | PHC/OPD attendees | F | Adult | 15 to 44 | Non-stratified | NG, TV |
| EA | Senkoro 2024[171] | 2022 | Tanzania | GYN attendees | F | Adult | 18 to 45 | Non-stratified | NG, TV |
| SA | Romoren 2007[172] | 2000 | Botswana | ANC attendees | F | Adult | 15 to 43 | Non-stratified | NG, TV |
| SA | Kleinschmidt 2007[173] | 2000 | South Africa | FP attendees | F | Adult | 18 to 40 | HIV negative | NG, TV |

*Continued...*

| Region | Reference | Year | Country | Population | Sex | Age | Age range | HIV status | Infections |
|---|---|---|---|---|---|---|---|---|---|
| SA | Sturm 2004[174] | 2001* | South Africa | ANC attendees | F | Adult | NR | Non-stratified | CT, NG, TV |
| SA | Paz-Bailey 2005[175] | 2002 | Botswana | FP attendees | F | Adult | NR | Non-stratified | NG, TV |
| SA | Pettifor 2005[176] | 2002 | South Africa | PHC/OPD attendees | F, M | Youth | 15 to 24 | Non-stratified | CT, NG |
| SA | van de Wijgert 2006[177] | 2002 | South Africa | PHC/OPD attendees | F | Adult | 18 to 69 | Non-stratified | CT, NG, TV |
| SA | Odendaal 2006[178] | 2003 | South Africa | ANC attendees | F | Adult | NR | Non-stratified | CT, NG |
| SA | Sobngwi-Tambekou 2009[179] | 2003 | South Africa | Prevention trial participants | M | Adult | 18 to 24 | Non-stratified | CT, NG, TV |
| SA | Ramjee 2008[99] | 2004 | South Africa | Prevention trial participants | F | Adult | 18+ | Non-stratified | CT, NG, TV |
| SA | Sebitloane 2011[180] | 2004 | South Africa | ANC attendees | F | Adult | 18+ | HIV negative | TV |
| SA | Venkatesh 2011[100] | 2004 | South Africa | Prevention trial participants | F | Adult | 18 to 49 | HIV negative | CT, NG, TV |
| SA | Black 2008[181] | 2005 | South Africa | PHC/OPD attendees | M | Adult | NR | Non-stratified | CT, NG, TV |
| SA | Guffey 2014[104] | 2006 | South Africa | Prevention trial participants | F | Adult | 18+ | HIV negative | CT, NG, TV |
| SA | Lewis 2008[182] | 2006 | South Africa | PHC/OPD attendees | M | Adult | 17 to 73 | Non-stratified | CT, NG, TV |
| SA | De Jongh 2010[183] | 2007* | South Africa | GYN attendees | F | Adult | 13 to 41 | Non-stratified | CT, NG, TV |
| SA | McCormack 2010[105] | 2007 | South Africa | Prevention trial participants | F | Adult | 18+ | HIV negative | CT, NG, TV |
| SA | Botswana Ministry of Health 2011[184] | 2008 | Botswana | FP attendees | F | Adult | NR | Non-stratified | CT, NG |
| SA | Thigpen 2012[185] | 2008 | Botswana | Prevention trial participants | F | Adult | 18 to 39 | HIV negative | TV |
| SA | Moodley 2015[186] | 2009 | South Africa | ANC attendees | F | Adult | 18+ | Non-stratified | CT, NG, TV |
| SA | Jespers 2014[115] | 2011 | South Africa | ANC attendees, FP attendees | F | Adult | 18 to 35 | Non-stratified | CT, NG, TV |
| SA | Kleppa 2015[187] | 2011 | South Africa | Students | F | Youth | 15 to 31 | Non-stratified | CT, NG, TV |
| SA | Peters 2014[188] | 2011 | South Africa | PHC/OPD attendees | F | Adult | 18 to 49 | Non-stratified | CT, NG |
| SA | Galappaththi-Arachchige 2016[189] | 2012 | South Africa | Students | F | Youth | 16 to 20 | Non-stratified | CT, NG, TV |
| SA | Shukla 2023[190] | 2012 | South Africa | Students | F | Youth | 16 to 22 | Non-stratified | CT, NG, TV |
| SA | Jewanraj 2021[191] | 2013 | South Africa | Prevention trial participants | F | Adult | 20 to 44 | HIV negative | CT, NG, TV |
| SA | Jongen 2021[192] | 2013 | South Africa | PHC/OPD attendees | F | Youth | 16 to 24 | HIV negative | CT, NG |
| SA | Palanee-Phillips 2015[129] | 2013 | South Africa | Prevention trial participants | F | Adult | 18 to 45 | HIV negative | CT, NG, TV |

*Continued...*

| Region | Reference | Year | Country | Population | Sex | Age | Age range | HIV status | Infections |
|---|---|---|---|---|---|---|---|---|---|
| SA | Barnabas 2018[193] | 2014 | South Africa | PHC/OPD attendees | F | Youth | 16 to 22 | HIV negative | CT, NG, TV |
| SA | Le Roux 2017[194] | 2014* | South Africa | PHC/OPD attendees | M | Adult | 17 to 65 | Non-stratified | CT, NG, TV |
| SA | Ginindza 2017[195] | 2015 | Eswatini | PHC/OPD attendees | F | Adult | 15 to 49 | Non-stratified | CT, NG, TV |
| SA | Abbai-Shaik 2016[196] | 2015 | South Africa | PHC/OPD attendees | M | Adult | 18+ | Non-stratified | CT |
| SA | Huyveneers 2023[197] | 2015 | South Africa | Prevention trial participants | F | Adult | 18 to 45 | HIV negative | CT, NG, TV |
| SA | Kaida 2018[198] | 2015 | South Africa | Community members | F, M | Youth | 16 to 24 | Non-stratified | CT, NG, TV |
| SA | Kharsany 2020[199] | 2015 | South Africa | Population-representative survey participants | F, M | Adult | 15 to 49 | Non-stratified | CT, NG, TV |
| SA | Wynn 2018[200] | 2016 | Botswana | ANC attendees | F | Adult | 18+ | Non-stratified | CT, NG, TV |
| SA | Deese 2021[147] | 2016 | Eswatini, South Africa | FP attendees | F | Adult | 16 to 35 | HIV negative | CT, NG |
| SA | Gorgens 2020[201] | 2016 | Eswatini | Prevention trial participants | F | Youth | 15 to 22 | HIV negative | TV |
| SA | Gill 2020[202] | 2016 | South Africa | Prevention trial participants | F | Youth | 15 to 19 | HIV negative | CT, NG, TV |
| SA | Hoffman 2019[203] | 2016 | South Africa | PHC/OPD attendees | F | Adult | 18 to 75 | Non-stratified | CT, NG, TV |
| SA | Mgodi 2021[148] | 2017 | Botswana, South Africa | Prevention trial participants | F | Adult | 18 to 50 | HIV negative | CT, NG |
| SA | Delany-Moretlwe 2023[204] | 2017 | South Africa, Zimbabwe | Prevention trial participants | F | Youth | 16 to 25 | HIV negative | CT, NG, TV |
| SA | Francis 2018[205] | 2017 | South Africa | Population-representative survey participants | F, M | Youth | 15 to 24 | Non-stratified | CT, NG, TV |
| SA | Dessai 2020[206] | 2018 | South Africa | ANC attendees | F | Adult | 18 to 43 | Non-stratified | TV |
| SA | Govender 2023[207] | 2018 | South Africa | ANC attendees | F | Adult | 15+ | HIV negative | CT, NG, TV |
| SA | Gray 2021[208] | 2018 | South Africa | Prevention trial participants | F, M | Adult | 18 to 35 | HIV negative | CT, NG, TV |
| SA | Joseph Davey 2019[209] | 2018 | South Africa | ANC attendees | F | Adult | 18+ | Non-stratified | CT, NG, TV |
| SA | Naicker 2021[210] | 2018 | South Africa | ANC attendees | F | Adult | 18+ | Non-stratified | TV |
| SA | Price 2024[211] | 2018 | South Africa | Community members | F, M | Youth | 12 to 19 | HIV negative | CT, NG |
| SA | Taku 2021[212] | 2018 | South Africa | PHC/OPD attendees | F | Adult | 30+ | Non-stratified | CT, NG, TV |
| SA | Celum 2022[155] | 2019 | South Africa | FP attendees | F | Youth | 16 to 25 | HIV negative | CT, NG |
| SA | Chetty 2020[213] | 2019 | South Africa | ANC attendees | F | Adult | 18+ | Non-stratified | TV |

*Continued...*

| Region | Reference | Year | Country | Population | Sex | Age | Age range | HIV status | Infections |
|---|---|---|---|---|---|---|---|---|---|
| SA | Oree 2021[214] | 2019 | South Africa | ANC attendees | F | Adult | 20 to 40 | Non-stratified | NG |
| SA | de Voux 2023[215] | 2020 | South Africa | ANC attendees | F | Adult | 16+ | HIV negative | CT, NG, TV |
| SA | Mabaso 2022[216] | 2020 | South Africa | ANC attendees | F | Adult | 18+ | Non-stratified | CT |
| SA | Nair 2023[164] | 2020 | South Africa | Prevention trial participants | F | Youth | 16 to 21 | HIV negative | CT, NG, TV |
| SA | Jarolimova 2023[217] | 2021 | South Africa | Population-representative survey participants | F, M | Youth | 16 to 29 | Non-stratified | CT, NG, TV |
| SA | Mullick 2023[218] | 2021 | South Africa | PHC/OPD attendees | F, M | Youth | 15 to 24 | HIV negative | CT, NG, TV |
| SA | Mussa 2023[219] | 2022 | Botswana | ANC attendees | F | Adult | 15+ | Non-stratified | CT, NG |
| SA | de Voux 2024[220] | 2022 | South Africa | ANC attendees | F | Adult | 18+ | Non-stratified | CT, NG, TV |
| Multiple | Delany-Moretlwe 2022[221] | 2019 | Botswana, Eswatini, Kenya, Malawi, South Africa, Uganda, Zimbabwe | Prevention trial participants | F | Adult | 18 to 45 | HIV negative | CT, NG, TV |

**Region** — CA: Central Africa, EA: Eastern Africa, SA: Southern Africa, WA: Western Africa.
**Year** — Midpoint year between start and end of data collection.*For studies without data collection dates reported, year was estimated by subtracting the median publication lag from the publication year (three years, based on difference among studies with dates reported).
**Population** — ANC: antenatal care, FP: family planning clinic, GYN: gynaecology clinic, PHC/OPD: primary healthcare or outpatient department.
**Sex** — F: Female, M: Male.
**Age** — Youth: 12–25 years, Adult: 12+ years.
**Infections** — CT: *Chlamydia trachomatis*, NG: *Neisseria gonorrhoeae*, TV: *Trichomonas vaginalis*.

The full study database is included as a separate supplementary file.

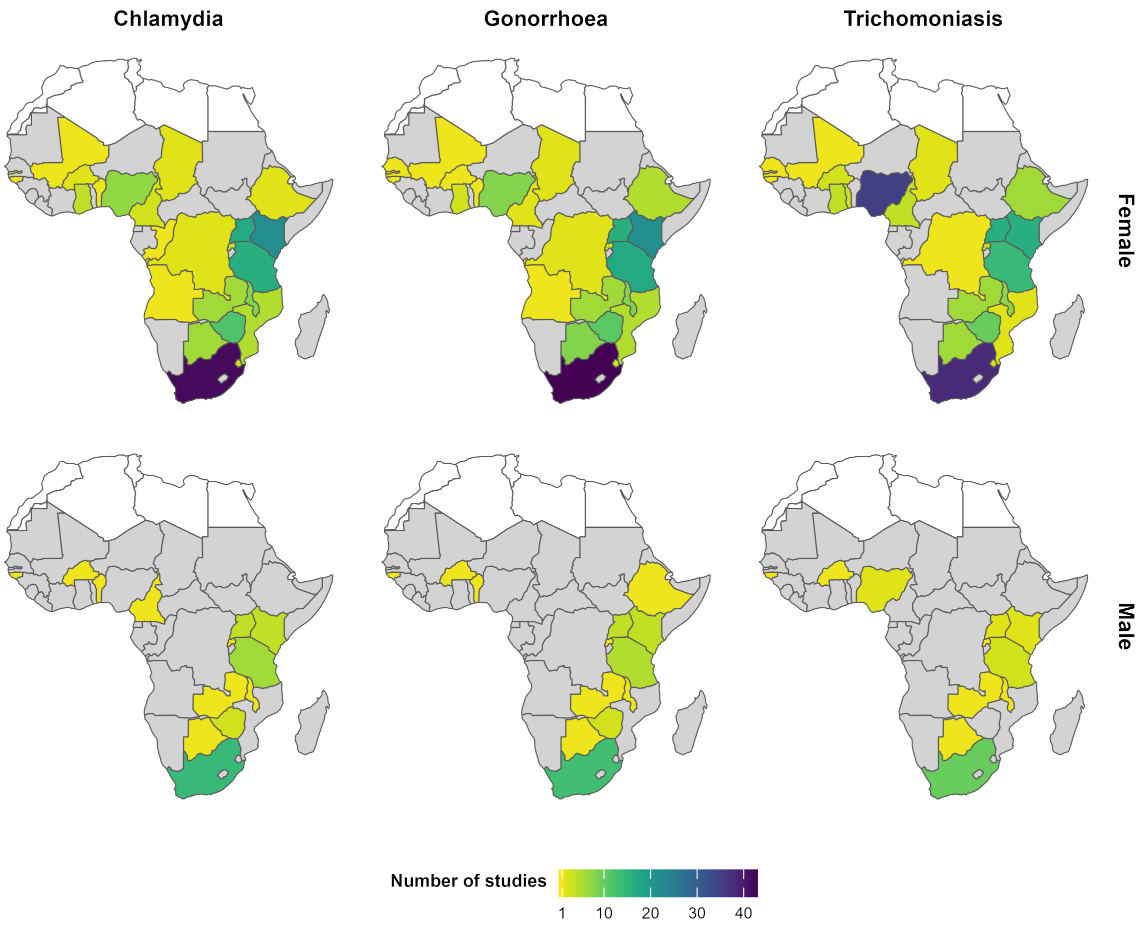

**Figure S1:** Number of studies included per country in sub-Saharan Africa.

Number of studies assessing the prevalence of chlamydia, gonorrhoea, and trichomoniasis among females and males in sub-Saharan Africa. Grey shading represents countries with no identified studies. White shading represents countries outside of sub-Saharan Africa, according to the UN M49 Standard.[1] Total included studies were 139 for chlamydia, 140 for gonorrhoea, and 162 for trichomoniasis. Source for base map data is Natural Earth.[222]

**Table S6:** Adjusted prevalence ratios for chlamydia, gonorrhoea, and trichomoniasis in sub-Saharan Africa, estimated via log-binomial generalised linear mixed-effects models: *within-study analysis using observations adjusted for diagnostic test performance*

| **Variable** | **Chlamydia aPR (95% CI)** | **Gonorrhoea aPR (95% CI)** | **Trichomoniasis aPR (95% CI)** |
|---|---|---|---|
| **Intercept** | 0.13 (0.08-0.21) | 0.04 (0.02-0.07) | 0.21 (0.17-0.26) |
| **Sub-region** | | | |
| Western and Central | 0.28 (0.13-0.61) | 0.54 (0.17-1.66) | 0.46 (0.28-0.74) |
| Eastern | 0.43 (0.27-0.68) | 0.52 (0.26-1.04) | 1.20 (0.86-1.67) |
| Southern | Ref | Ref | Ref |
| **Sub-region:year*** | | | |
| Western and Central:Year | 1.04 (0.96-1.12) | 1.18 (1.04-1.33) | 1.02 (1.00-1.04) |
| Eastern:Year | 1.07 (1.03-1.10) | 1.00 (0.95-1.05) | 1.06 (0.82-1.39) |
| Southern:Year | 1.05 (1.00-1.11) | 1.05 (0.97-1.14) | 0.89 (0.84-0.95) |
| **Sex** | | | |
| Female | Ref | Ref | Ref |
| Male | 0.61 (0.56-0.65) | 0.73 (0.65-0.83) | 0.23 (0.20-0.27) |
| **Age group** | | | |
| Adult | Ref | Ref | Ref |
| Youth | 1.17 (0.76-1.80) | 1.25 (0.66-2.34) | 0.36 (0.22-0.57) |
| **HIV status** | | | |
| Non-stratified | Ref | Ref | Ref |
| HIV negative | 1.08 (0.67-1.73) | 1.05 (0.52-2.14) | 0.73 (0.19-2.70) |
| **Diagnostic test** | | | |
| NAAT | Ref | Ref | Ref |
| Culture | - | 7.87 (1.92-32.19) | 2.48 (1.50-4.10) |
| DFA | 1.76 (0.52-5.94) | - | - |
| Rapid antigen test | - | - | 0.40 (0.05-3.40) |
| Wet mount | - | - | 0.88 (0.13-6.02) |
| **Model variance** | | | |
| $\tau^2$ fixed | 0.68 | 0.59 | 1.16 |
| $\tau^2$ random | 0.17 | 0.37 | <0.01 |
| **Number groups** | | | |
| Number studies | 26 | 26 | 12 |
| Number observations | 52 | 52 | 24 |

*Midpoint year between start and end of data collection period; centred at 2012. aPR: adjusted prevalence ratio, 95% CI: 95% confidence interval, DFA: direct fluorescent antibody, NAAT: nucleic acid amplification test, $\tau^2$: variance.

**Table S7:** Adjusted prevalence ratios for chlamydia, gonorrhoea, and trichomoniasis in sub-Saharan Africa, estimated via log-binomial generalised linear mixed-effects models: *between-study sensitivity analysis using observations unadjusted for diagnostic test performance*

| **Variable** | **Chlamydia aPR (95% CI)** | **Gonorrhoea aPR (95% CI)** | **Trichomoniasis aPR (95% CI)** |
|---|---|---|---|
| **Intercept** | 0.13 (0.10-0.16) | 0.03 (0.02-0.05) | 0.10 (0.07-0.13) |
| **Sub-region** | | | |
| Western and Central | 0.29 (0.21-0.40) | 0.32 (0.20-0.52) | 0.89 (0.64-1.23) |
| Eastern | 0.36 (0.34-0.40) | 0.63 (0.56-0.72) | 0.99 (0.85-1.17) |
| Southern | Ref | Ref | Ref |
| **Sub-region:year*** | | | |
| Western and Central:Year | 0.99 (0.95-1.02) | 0.99 (0.93-1.04) | 0.99 (0.96-1.02) |
| Eastern:Year | 1.07 (1.05-1.10) | 1.01 (0.99-1.04) | 0.97 (0.94-1.01) |
| Southern:Year | 1.01 (0.99-1.03) | 1.01 (0.98-1.04) | 0.96 (0.92-0.99) |
| **Sub-region:sex†** | | | |
| Western and Central:Male | 0.79 (0.57-1.08) | 0.86 (0.45-1.66) | 0.15 (0.10-0.23) |
| Eastern:Male | 0.73 (0.65-0.82) | 0.58 (0.48-0.70) | 0.53 (0.46-0.62) |
| Southern:Male | 0.64 (0.59-0.69) | 0.60 (0.51-0.69) | 0.26 (0.22-0.30) |
| **Population** | | | |
| ANC attendees | Ref | Ref | Ref |
| FP attendees | 0.97 (0.64-1.48) | 0.87 (0.46-1.66) | 0.50 (0.29-0.85) |
| GYN attendees | 0.97 (0.53-1.76) | 0.69 (0.31-1.56) | 1.47 (0.89-2.43) |
| PHC/OPD attendees | 0.77 (0.55-1.09) | 1.43 (0.86-2.35) | 0.85 (0.56-1.29) |
| Students | 1.05 (0.69-1.62) | 1.21 (0.58-2.56) | 1.12 (0.70-1.79) |
| Community members | 0.99 (0.67-1.46) | 1.14 (0.65-1.99) | 0.88 (0.46-1.68) |
| Prevention trial participants | 0.87 (0.60-1.26) | 1.10 (0.64-1.89) | 0.97 (0.60-1.55) |
| Population-representative survey participants | 0.92 (0.59-1.44) | 1.25 (0.65-2.40) | 1.29 (0.71-2.35) |
| **Age group** | | | |
| Adult | Ref | Ref | Ref |
| Youth | 1.12 (0.84-1.49) | 1.01 (0.66-1.54) | 0.63 (0.40-0.99) |
| **HIV status** | | | |
| Non-stratified | Ref | Ref | Ref |
| HIV negative | 1.45 (1.09-1.94) | 1.21 (0.79-1.85) | 0.82 (0.57-1.17) |
| **Diagnostic test** | | | |
| NAAT | Ref | Ref | Ref |
| Culture | - | 1.04 (0.63-1.71) | 0.84 (0.55-1.28) |
| DFA | 0.78 (0.37-1.64) | - | - |
| ELISA | 1.55 (0.58-4.12) | - | - |
| Rapid antigen test | 1.92 (1.16-3.18) | 1.95 (0.35-10.89) | 0.87 (0.48-1.58) |
| Wet mount | - | - | 0.74 (0.53-1.04) |
| **Model variance** | | | |
| $\tau^2$ fixed | 0.47 | 0.22 | 0.22 |
| $\tau^2$ random | 0.34 | 0.69 | 0.59 |
| **Number groups** | | | |
| Number studies | 139 | 140 | 162 |
| Number observations | 202 | 202 | 207 |

*Midpoint year between start and end of data collection period; centred at 2012. †Reference sex is female. aPR: adjusted prevalence ratio, 95% CI: 95% confidence interval, DFA: direct fluorescent antibody, ELISA: enzyme-linked immunosorbent assay, NAAT: nucleic acid amplification test, $\tau^2$: variance.

**Table S8:** Adjusted prevalence ratios for chlamydia, gonorrhoea, and trichomoniasis in sub-Saharan Africa, estimated via log-binomial generalised linear mixed-effects models: *between-study sensitivity analysis using NAAT-diagnosed observations adjusted for test performance*

| **Variable** | **Chlamydia aPR (95% CI)** | **Gonorrhoea aPR (95% CI)** | **Trichomoniasis aPR (95% CI)** |
|---|---|---|---|
| **Intercept** | 0.15 (0.12-0.18) | 0.04 (0.03-0.06) | 0.14 (0.09-0.23) |
| **Sub-region** | | | |
| Western and Central | 0.29 (0.21-0.40) | 0.28 (0.17-0.47) | 0.99 (0.40-2.46) |
| Eastern | 0.38 (0.35-0.41) | 0.64 (0.56-0.73) | 0.58 (0.36-0.93) |
| Southern | Ref | Ref | Ref |
| **Sub-region:year*** | | | |
| Western and Central:Year | 1.01 (0.97-1.05) | 1.06 (0.99-1.13) | 0.88 (0.75-1.03) |
| Eastern:Year | 1.07 (1.05-1.09) | 1.02 (0.99-1.04) | 0.95 (0.88-1.02) |
| Southern:Year | 1.02 (1.00-1.04) | 1.02 (0.99-1.05) | 0.96 (0.90-1.02) |
| **Sub-region:sex†** | | | |
| Western and Central:Male | 0.55 (0.33-0.91) | 1.13 (0.57-2.21) | 0.04 (0.01-0.33) |
| Eastern:Male | 0.61 (0.53-0.70) | 0.70 (0.56-0.86) | 0.39 (0.31-0.47) |
| Southern:Male | 0.61 (0.56-0.67) | 0.80 (0.69-0.94) | 0.15 (0.11-0.20) |
| **Population** | | | |
| ANC attendees | Ref | Ref | Ref |
| FP attendees | 0.94 (0.61-1.45) | 0.93 (0.50-1.73) | 0.06 (0.00-0.82) |
| GYN attendees | 0.98 (0.56-1.71) | 0.65 (0.27-1.56) | 0.69 (0.28-1.70) |
| PHC/OPD attendees | 0.79 (0.57-1.11) | 1.25 (0.77-2.03) | 0.97 (0.48-1.93) |
| Students | 0.84 (0.54-1.30) | 0.92 (0.48-1.76) | 2.24 (1.05-4.77) |
| Community members | 0.93 (0.64-1.35) | 0.82 (0.48-1.39) | 0.68 (0.23-2.02) |
| Prevention trial participants | 0.84 (0.59-1.19) | 1.07 (0.65-1.73) | 0.95 (0.39-2.31) |
| Population-representative survey participants | 0.91 (0.60-1.40) | 1.13 (0.63-2.02) | 0.79 (0.31-1.99) |
| **Age group** | | | |
| Adult | Ref | Ref | Ref |
| Youth | 1.22 (0.93-1.62) | 1.24 (0.85-1.79) | 0.33 (0.16-0.69) |
| **HIV status** | | | |
| Non-stratified | Ref | Ref | Ref |
| HIV negative | 1.42 (1.07-1.88) | 0.96 (0.65-1.42) | 0.62 (0.32-1.22) |
| **Model variance** | | | |
| $\tau^2$ fixed | 0.53 | 0.24 | 1.07 |
| $\tau^2$ random | 0.29 | 0.48 | 0.65 |
| **Number groups** | | | |
| Number studies | 127 | 115 | 56 |
| Number observations | 188 | 176 | 68 |

*Midpoint year between start and end of data collection period; centred at 2012. †Reference sex is female. aPR: adjusted prevalence ratio, 95% CI: 95% confidence interval, NAAT: nucleic acid amplification test, $\tau^2$: variance.

**Table S9:** Adjusted prevalence ratios for chlamydia, gonorrhoea, and trichomoniasis in sub-Saharan Africa, estimated via log-binomial generalised linear mixed-effects models: *between-study sensitivity analysis using NAAT-diagnosed observations unadjusted for test performance*

| **Variable** | **Chlamydia aPR (95% CI)** | **Gonorrhoea aPR (95% CI)** | **Trichomoniasis aPR (95% CI)** |
|---|---|---|---|
| **Intercept** | 0.13 (0.11-0.16) | 0.04 (0.03-0.06) | 0.16 (0.11-0.24) |
| **Sub-region** | | | |
| Western and Central | 0.27 (0.20-0.38) | 0.28 (0.17-0.47) | 0.98 (0.45-2.14) |
| Eastern | 0.37 (0.34-0.40) | 0.63 (0.56-0.71) | 0.64 (0.44-0.94) |
| Southern | Ref | Ref | Ref |
| **Sub-region:year*** | | | |
| Western and Central:Year | 1.01 (0.97-1.05) | 1.05 (0.99-1.12) | 0.89 (0.78-1.02) |
| Eastern:Year | 1.07 (1.05-1.10) | 1.02 (0.99-1.05) | 0.94 (0.89-1.00) |
| Southern:Year | 1.01 (0.99-1.03) | 1.02 (0.99-1.05) | 0.96 (0.92-1.01) |
| **Sub-region:sex†** | | | |
| Western and Central:Male | 0.74 (0.50-1.10) | 0.90 (0.46-1.74) | 0.00 (0.00-Inf) |
| Eastern:Male | 0.73 (0.65-0.82) | 0.57 (0.47-0.69) | 0.58 (0.49-0.67) |
| Southern:Male | 0.64 (0.59-0.69) | 0.62 (0.54-0.72) | 0.26 (0.22-0.30) |
| **Population** | | | |
| ANC attendees | Ref | Ref | Ref |
| FP attendees | 0.98 (0.64-1.51) | 0.97 (0.51-1.85) | 0.19 (0.03-1.08) |
| GYN attendees | 0.92 (0.52-1.61) | 0.50 (0.20-1.27) | 0.64 (0.29-1.40) |
| PHC/OPD attendees | 0.76 (0.54-1.06) | 1.15 (0.70-1.90) | 0.83 (0.46-1.50) |
| Students | 0.84 (0.55-1.31) | 0.95 (0.48-1.88) | 1.71 (0.90-3.25) |
| Community members | 0.95 (0.66-1.37) | 0.83 (0.48-1.43) | 0.49 (0.19-1.23) |
| Prevention trial participants | 0.83 (0.58-1.17) | 0.99 (0.60-1.65) | 0.82 (0.39-1.74) |
| Population-representative survey participants | 0.93 (0.61-1.42) | 1.14 (0.62-2.08) | 0.71 (0.32-1.56) |
| **Age group** | | | |
| Adult | Ref | Ref | Ref |
| Youth | 1.21 (0.92-1.59) | 1.14 (0.78-1.68) | 0.44 (0.24-0.82) |
| **HIV status** | | | |
| Non-stratified | Ref | Ref | Ref |
| HIV negative | 1.45 (1.10-1.92) | 1.07 (0.72-1.60) | 0.64 (0.36-1.14) |
| **Model variance** | | | |
| $\tau^2$ fixed | 0.51 | 0.26 | 6.56 |
| $\tau^2$ random | 0.29 | 0.54 | 0.48 |
| **Number groups** | | | |
| Number studies | 127 | 115 | 56 |
| Number observations | 188 | 176 | 68 |

*Midpoint year between start and end of data collection period; centred at 2012. †Reference sex is female. aPR: adjusted prevalence ratio, 95% CI: 95% confidence interval, NAAT: nucleic acid amplification test, $\tau^2$: variance.

**Table S10:** Adjusted prevalence ratios for chlamydia, gonorrhoea, and trichomoniasis in sub-Saharan Africa, estimated via log-binomial generalised linear mixed-effects models: *within-study sensitivity analysis using observations unadjusted for diagnostic test performance*

| **Variable** | **Chlamydia aPR (95% CI)** | **Gonorrhoea aPR (95% CI)** | **Trichomoniasis aPR (95% CI)** |
|---|---|---|---|
| **Intercept** | 0.12 (0.08-0.19) | 0.04 (0.02-0.08) | 0.22 (0.18-0.25) |
| **Sub-region** | | | |
| Western and Central | 0.28 (0.13-0.57) | 0.45 (0.15-1.35) | 0.35 (0.23-0.52) |
| Eastern | 0.44 (0.28-0.67) | 0.48 (0.24-0.95) | 0.80 (0.63-1.02) |
| Southern | Ref | Ref | Ref |
| **Sub-region:year*** | | | |
| Western and Central:Year | 1.03 (0.95-1.11) | 1.19 (1.05-1.35) | 1.02 (1.00-1.04) |
| Eastern:Year | 1.07 (1.03-1.10) | 1.00 (0.95-1.05) | 0.95 (0.80-1.12) |
| Southern:Year | 1.05 (1.00-1.10) | 1.04 (0.96-1.13) | 0.90 (0.86-0.94) |
| **Sex** | | | |
| Female | Ref | Ref | Ref |
| Male | 0.67 (0.63-0.71) | 0.59 (0.52-0.66) | 0.34 (0.31-0.38) |
| **Age group** | | | |
| Adult | Ref | Ref | Ref |
| Youth | 1.08 (0.72-1.62) | 1.04 (0.56-1.93) | 0.38 (0.27-0.54) |
| **HIV status** | | | |
| Non-stratified | Ref | Ref | Ref |
| HIV negative | 1.14 (0.73-1.77) | 1.17 (0.59-2.32) | 0.60 (0.26-1.40) |
| **Diagnostic test** | | | |
| NAAT | Ref | Ref | Ref |
| Culture | - | 6.18 (1.55-24.62) | 2.07 (1.31-3.25) |
| DFA | 1.71 (0.55-5.31) | - | - |
| Rapid antigen test | - | - | 1.13 (0.28-4.62) |
| Wet mount | - | - | 0.00 (0.00-Inf) |
| **Model variance** | | | |
| $\tau^2$ fixed | 0.61 | 0.66 | 33.86 |
| $\tau^2$ random | 0.15 | 0.35 | <0.01 |
| **Number groups** | | | |
| Number studies | 26 | 26 | 12 |
| Number observations | 52 | 52 | 24 |

*Midpoint year between start and end of data collection period; centred at 2012. aPR: adjusted prevalence ratio, 95% CI: 95% confidence interval, DFA: direct fluorescent antibody, NAAT: nucleic acid amplification test, $\tau^2$: variance.

**Table S11:** Adjusted prevalence ratios for chlamydia, gonorrhoea, and trichomoniasis in sub-Saharan Africa, estimated via log-binomial generalised linear mixed-effects models: *within-study sensitivity analysis using NAAT-diagnosed observations adjusted for test performance*

| **Variable** | **Chlamydia aPR (95% CI)** | **Gonorrhoea aPR (95% CI)** | **Trichomoniasis aPR (95% CI)** |
|---|---|---|---|
| **Intercept** | 0.13 (0.08-0.21) | 0.04 (0.02-0.08) | 0.21 (0.17-0.26) |
| **Sub-region** | | | |
| Western and Central | 0.28 (0.12-0.62) | 0.54 (0.17-1.70) | 0.48 (0.30-0.77) |
| Eastern | 0.43 (0.27-0.69) | 0.52 (0.25-1.05) | 1.18 (0.85-1.65) |
| Southern | Ref | Ref | Ref |
| **Sub-region:year*** | | | |
| Western and Central:Year | 1.04 (0.95-1.12) | 1.18 (1.04-1.33) | - |
| Eastern:Year | 1.07 (1.03-1.10) | 1.00 (0.95-1.05) | 1.07 (0.82-1.40) |
| Southern:Year | 1.05 (1.00-1.11) | 1.05 (0.97-1.14) | 0.89 (0.84-0.95) |
| **Sex** | | | |
| Female | Ref | Ref | Ref |
| Male | 0.60 (0.56-0.65) | 0.74 (0.65-0.84) | 0.25 (0.22-0.30) |
| **Age group** | | | |
| Adult | Ref | Ref | Ref |
| Youth | 1.17 (0.75-1.81) | 1.24 (0.65-2.36) | 0.35 (0.22-0.56) |
| **HIV status** | | | |
| Non-stratified | Ref | Ref | Ref |
| HIV negative | 1.08 (0.66-1.75) | 1.05 (0.51-2.18) | 0.74 (0.20-2.76) |
| **Model variance** | | | |
| $\tau^2$ fixed | 0.70 | 0.49 | 1.09 |
| $\tau^2$ random | 0.17 | 0.39 | <0.01 |
| **Number groups** | | | |
| Number studies | 25 | 25 | 8 |
| Number observations | 50 | 50 | 16 |

*Midpoint year between start and end of data collection period; centred at 2012. †Reference sex is female. aPR: adjusted prevalence ratio, 95% CI: 95% confidence interval, NAAT: nucleic acid amplification test, $\tau^2$: variance.

**Table S12:** Adjusted prevalence ratios for chlamydia, gonorrhoea, and trichomoniasis in sub-Saharan Africa, estimated via log-binomial generalised linear mixed-effects models: *within-study sensitivity analysis using NAAT-diagnosed observations unadjusted for test performance*

| **Variable** | **Chlamydia aPR (95% CI)** | **Gonorrhoea aPR (95% CI)** | **Trichomoniasis aPR (95% CI)** |
|---|---|---|---|
| **Intercept** | 0.12 (0.08-0.19) | 0.04 (0.02-0.08) | 0.21 (0.18-0.25) |
| **Sub-region** | | | |
| Western and Central | 0.28 (0.13-0.58) | 0.45 (0.15-1.37) | 0.37 (0.25-0.55) |
| Eastern | 0.43 (0.28-0.68) | 0.48 (0.24-0.96) | 0.80 (0.63-1.01) |
| Southern | Ref | Ref | Ref |
| **Sub-region:year*** | | | |
| Western and Central:Year | 1.03 (0.95-1.11) | 1.19 (1.05-1.35) | - |
| Eastern:Year | 1.07 (1.03-1.10) | 1.00 (0.95-1.05) | 0.95 (0.80-1.13) |
| Southern:Year | 1.05 (0.99-1.10) | 1.04 (0.96-1.13) | 0.90 (0.86-0.94) |
| **Sex** | | | |
| Female | Ref | Ref | Ref |
| Male | 0.66 (0.62-0.71) | 0.59 (0.52-0.66) | 0.37 (0.33-0.41) |
| **Age group** | | | |
| Adult | Ref | Ref | Ref |
| Youth | 1.08 (0.72-1.63) | 1.04 (0.56-1.95) | 0.38 (0.26-0.54) |
| **HIV status** | | | |
| Non-stratified | Ref | Ref | Ref |
| HIV negative | 1.14 (0.72-1.79) | 1.17 (0.58-2.35) | 0.61 (0.26-1.43) |
| **Model variance** | | | |
| $\tau^2$ fixed | 0.63 | 0.58 | 0.79 |
| $\tau^2$ random | 0.15 | 0.36 | <0.01 |
| **Number groups** | | | |
| Number studies | 25 | 25 | 8 |
| Number observations | 50 | 50 | 16 |

*Midpoint year between start and end of data collection period; centred at 2012. †Reference sex is female. aPR: adjusted prevalence ratio, 95% CI: 95% confidence interval, NAAT: nucleic acid amplification test, $\tau^2$: variance.

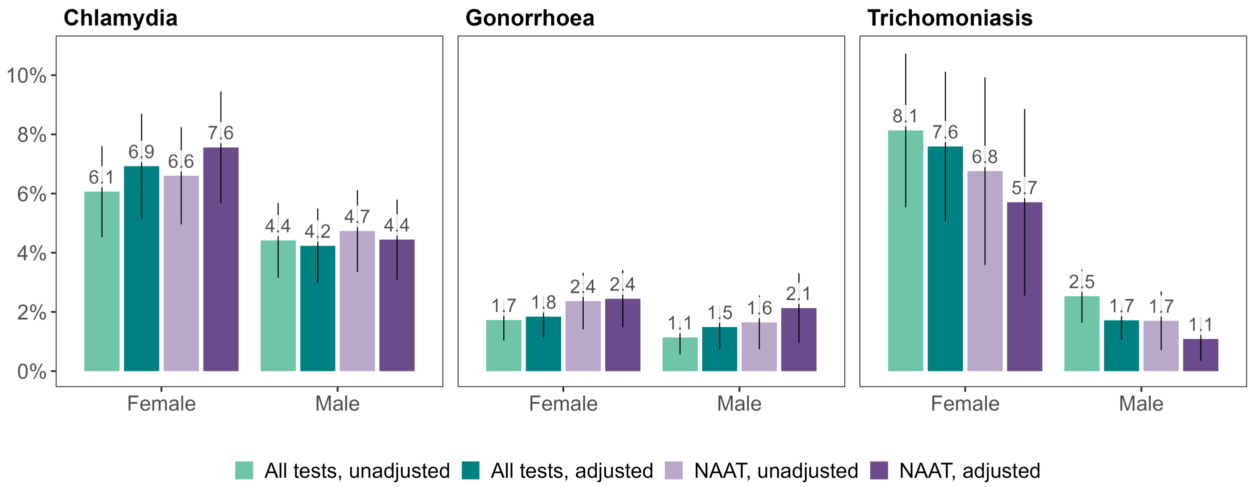

**Figure S2:** Sexually transmitted infection prevalence in sub-Saharan Africa in 2020, with and without accounting for diagnostic test performance.

Estimates of chlamydia, gonorrhoea, and trichomoniasis prevalence by sex for sub-Saharan Africa in 2020. Sub-regional estimates generated using log-binomial generalised linear mixed-effects models for each infection, using either observations as reported or adjusted for diagnostic test performance, with all diagnostic tests or NAAT only. Sub-Saharan African estimates represent sex-matched population-weighted means. Bars and error lines depict mean prevalence estimates with 95% confidence intervals. NAAT: Nucleic acid amplification test.

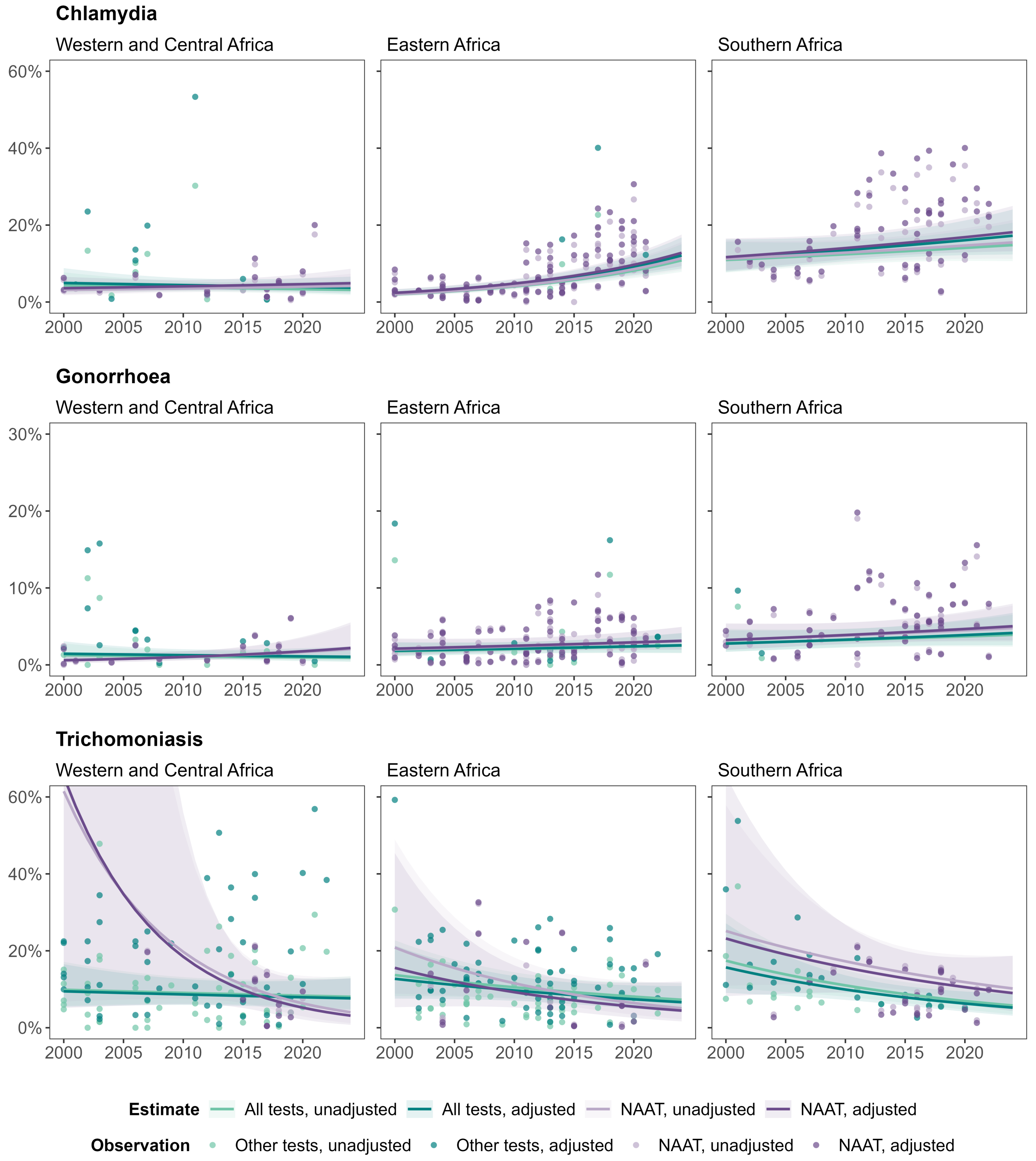

**Figure S3:** Sexually transmitted infection prevalence among females in sub-Saharan Africa between 2000 and 2024, with and without accounting for diagnostic test performance.

Estimates of chlamydia, gonorrhoea, and trichomoniasis prevalence among females between 2000 and 2024. Sub-regional estimates generated using log-binomial generalised linear mixed-effects models for each infection, using either observations as reported or adjusted for diagnostic test performance, with all diagnostic tests or NAAT only. Lines and shading depict mean prevalence estimates with 95% confidence intervals. Points represent study observations. NAAT: Nucleic acid amplification test.

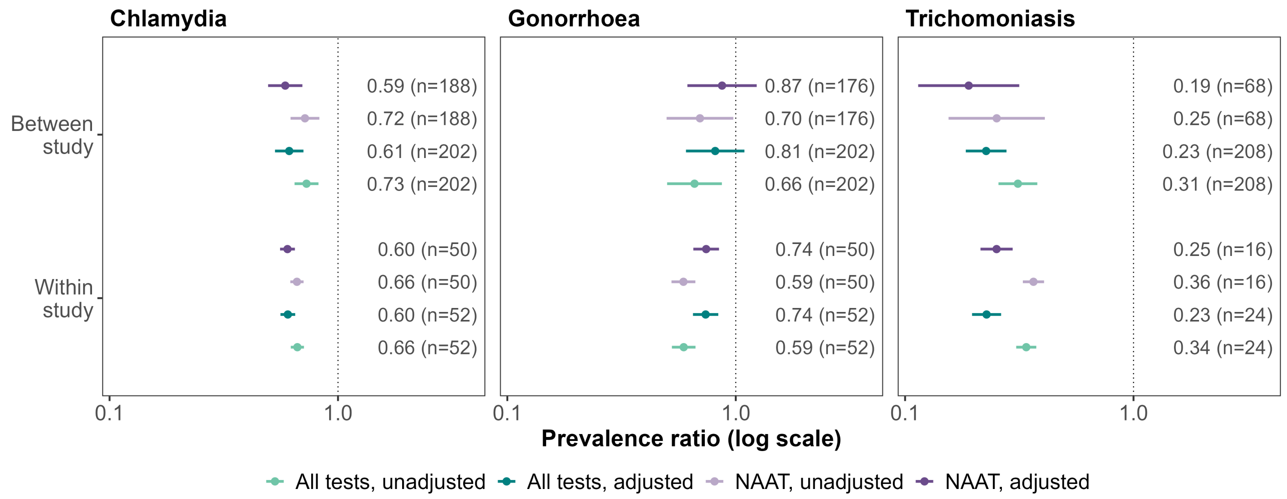

**Figure S4:** Sexually transmitted infection male-to-female prevalence ratio estimates in sub-Saharan Africa, with and without accounting for diagnostic test performance.

Male-to-female prevalence ratios for chlamydia, gonorrhoea, and trichomoniasis. Ratios estimated using log-binomial generalized linear mixed-effects models per infection, using either observations as reported or adjusted for diagnostic test performance, with all diagnostic tests or NAAT only. Models for within-study ratios used study observations among both sexes and between-study ratios used all study observations. Points and error lines depict population-weighted mean ratios and 95% confidence intervals for sub-Saharan Africa in 2020.

2. World Health Organization. Laboratory tests for the detection of reproductive tract infections. Geneva; 1999.

3. World Health Organization. Prevalence and incidence of selected sexually transmitted infections: Methods and results used by WHO to generate 2005 estimates. Geneva; 2011.

4. Grillo-Ardila CF, Torres M, Gaitán HG. Rapid point of care test for detecting urogenital Chlamydia trachomatis infection in nonpregnant women and men at reproductive age. The Cochrane database of systematic reviews. 2020 1;1(1).

5. Zhou Y, Jiang TT, Li J, Yin YP, Chen XS. Performance of point-of-care tests for the detection of chlamydia trachomatis infections: A systematic review and meta-analysis. EClinicalMedicine. 2021 7;37.

17. Tukur J, Fwacs M, Shittu SO, Drh MF, Abdul AM. A case control study of active genital Chlamydia trachomatis infection among patients with tubal infertility in northern Nigeria; 2006.

18. Balaka B, Agbèrè A, Dagnra A, Baeta S, Kessie K, Assimadi K. Genital bacterial carriage during the last trimester of pregnancy and early-onset neonatal sepsis. Archives de pédiatrie: organe officiel de la Sociéte française de pédiatrie. 2005;12(5):514–519.

19. Kirakoya-Samadoulogou F, Nagot N, Defer M, Yaro S, Meda N, Robert A. Bacterial Vaginosis Among Pregnant Women in Burkina Faso. Sexually transmitted diseases. 2008;35(12):985–989.

20. Siemer J, Theile O, Larbi Y, Fasching PA, Danso KA, Kreienberg R, et al. Chlamydia trachomatis Infection as a Risk Factor for Infertility among Women in Ghana, West Africa; 2008.

21. Chigbu LN, Aluka C, Eke RA. Trichomoniasis as an Indicator for Existing Sexually Transmitted Infections in Women in Aba, Nigeria; 2006. 1.

22. Inabo HI, Adewumi AA, Ishaya A. Prevalence of T. vaginalis in pregnant women attending antenatal clinics in Kaduna, Nigeria. Global Journal of Pure and Applied Sciences. 2006;12(3):323-5.

23. Jatau ED, Olonitola OS, Olayinka AT. Prevalence of Trichomonas Infection among Women Attending Antenatal Clinics in; 2006. 4.

49. Alexander SI, Idowu ET, Otubanjo OA, Ajayi MB. Trichomoniasis among pregnant women in Ifako Ijaiye, Shomolu
}and Agege Local Government Areas of Lagos State, Nigeria. Nigerian Journal of Parasitology. 2018;39(1):96-100.

72. Alexandre I, Justel M, Martinez P, Ortiz De Lejarazu R, Pastor JC. First attempt to implement ophthalmia neonatorum prophylaxis in Angola: Microorganisms, efficacy, and obstacles. Journal of Ophthalmology. 2015;2015.

78. Payne VK, Florence Cécile TT, Cedric Y, Christelle Nadia NA, José O. Risk Factors Associated with Prevalence of Candida albicans, Gardnerella vaginalis, and Trichomonas vaginalis among Women at the District Hospital of Dschang, West Region, Cameroon. International Journal of Microbiology. 2020;2020.

95. Mensch BS, Hewett PC, Helleringer S. Sexual Behavior and STI/HIV Status Among Adolescents in Rural Malawi: An Evaluation of the Effect of Interview Mode on Reporting NIH Public Access; 2008. 4.

160. Husen O, Aliyo A, Boru K, Gemechu T, Dedecha W, Ashenafi G. Trichomonas vaginalis and Associated Factors among Pregnant Women Attending Antenatal Care at Bule Hora University Teaching Hospital, Oromia Region, Southern Ethiopia. Journal of Parasitology Research. 2023;2023.

161. Zenebe MH, Mekonnen Z, Loha E, Padalko E. Prevalence, risk factors and association with delivery outcome of curable sexually transmitted infections among pregnant women in Southern Ethiopia. PLoS ONE. 2021 3;16(3 March).

172. Romoren M, Sundby J, Velauthapillai M, Rahman M, Klouman E, Hjortdahl P. Chlamydia and gonorrhoea in pregnant Batswana women: Time to discard the syndromic approach? BMC Infectious Diseases. 2007 4;7.
